# Defining the Global Landscape of Kidney Genetics Care— A Scoping Review and International Stakeholder Consultation of Clinic Models and Outcomes

**DOI:** 10.64898/2026.02.13.26346222

**Authors:** Ru Sin Lim, Trudie Harris, Julia Jefferis, Sadia Jahan, Regina Shaoying Lim, Louisa M D’Arrietta, Kar Hui Ng, Hui-Lin Chin, Liuh Ling Goh, Sanchalika Acharyya, Edwin Chan Chih-Yen, Chirag Patel, Erik Biros, Nick Sevdalis, Andrew J. Mallett, the Global Kidney Genetics Clinic Leads Collaborators

**Affiliations:** Department of Renal Medicine, Tan Tock Seng Hospital, Singapore; College of Medicine and Dentistry, James Cook University, Townsville, Queensland, Australia; Department of Renal Medicine, Townsville University Hospital, Townsville, Queensland, Australia; Department of Nephrology, Mater Health Service, Brisbane, Australia; Central and Northern Adelaide Renal and Transplantation Services, Royal Adelaide Hospital, Adelaide, SA, Australia; Library Services, Townsville University Hospital, Townsville Hospital and Health Service, Townsville, Queensland, Australia; Department of Paediatrics, Yong Loo Lin School of Medicine, National University of Singapore, Singapore; Khoo Teck Puat-National University Children’s Medical Institute, National University Health System, Singapore; Molecular Diagnostic Laboratory, Tan Tock Seng Hospital, Singapore; Clinical Research & Innovation Office, Tan Tock Seng Hospital, Singapore; Singapore Clinical Research Institute, Singapore; Genetic Health Queensland, Royal Brisbane and Women’s Hospital, Brisbane, Queensland, Australia; Faculty of Health, Medicine and Behavioural Sciences, The University of Queensland, Brisbane, Queensland, Australia; Centre for Behavioural and Implementation Science Interventions, Yong Loo Lin School of Medicine, National University of Singapore, Singapore; Faculty of Medicine and Institute for Molecular Bioscience, The University of Queensland, Brisbane, Queensland, Australia

**Author notes:** **Corresponding Author:** Ru Sin Lim, Department of Renal Medicine, Tan Tock Seng Hospital, 11 Jalan Tan Tock Seng, Singapore 308433, and Andrew J Mallett, Department of Renal Medicine, Townsville University Hospital, Angus Smith Drive, Dougla, QLD, 4814, Australia. The last two authors contributed equally and share senior authorship. The Global Kidney Genetics Clinic Leads Collaborators are listed in the Supplementary Appendix.

**Keywords:** Kidney Genetics Clinic Model, Genetic Kidney Disease, Nephrogenetics, Global Kidney Care, Scoping Review

## Abstract

**Introduction:** Genomic testing is reshaping nephrology practice, yet the structure, outcomes, and implementation of kidney genetics services remain poorly characterized.

**Methods:** We conducted a two-part scoping study comprising (i) a literature review (JBI methodology, PRISMA-ScR compliant; OSF registration <u>doi.org/10.17605/OSF.IO/N32VA</u>) of English-language publications (2000–2025) describing kidney genetics services and outcomes, and (ii) an international stakeholder consultation of clinic leads to capture real-world service and implementation experiences.

**Results:** Sixty studies were included, predominantly from North America (n=23), followed by United Kingdom/Ireland (n=5), Europe (n=17), Australia/New Zealand (n=10), and Asia (n=5). Among the 25 studies describing clinic models, four types were identified: multidisciplinary integrated (n=12), nephrologist-led (n=9), mainstreaming (n=2), and traditional genetics referral (n=2). Clinic structure varied by region. Outcome reporting focused on diagnostic yield (92%), with limited data on timeliness (16%), patient-reported outcomes (12%), or implementation outcomes (4%). Test penetration was high across regions and models, while diagnostic yield varied. Nephrologist-led clinics demonstrated comparable performance to multidisciplinary models when adequately supported.

International stakeholder consultation data (n=48) revealed diversification of clinic models across regions. In Australia/New Zealand, multidisciplinary clinics predominated, supported by public funding and in-house or hybrid laboratory. United Kingdom/Ireland clinics used public funding and a national laboratory. North American clinics show greater heterogeneity, with higher prevalence of nephrologist-led models, reliance on commercial laboratories, and mixed or private funding. Asian clinics reported nephrologist-led models, with resource constraints, and hybrid testing and funding arrangements. Comprehensive sequencing with virtual panels predominated in Australia/New Zealand, United Kingdom, and Europe; phenotype-driven panels ± reflex testing were more common in North America.

**Conclusions:** Kidney genetics care is expanding but remains unevenly implemented. Nephrologist-led and multidisciplinary models can be effective with appropriate support. Patient selection may influence diagnostic yield more than testing modality. Standardized outcome reporting and theory-driven implementation evaluation are essential for delivering equitable, sustainable genomic services.

**Lay Summary:** This study examined how kidney genetics services are delivered across the globe. We reviewed 60 studies (2000–2025) and consulted 48 clinic leaders globally. Four service models were identified—multidisciplinary integrated, nephrologist-led, mainstreaming, and traditional genetics referral—and mapped variation in care teams, test strategies, test laboratories, and funding. Most studies reported diagnostic yield, but few assessed patient experience or how well services were implemented. European programs showed the highest performance, attributed to clear referral criteria, deep phenotyping, detailed family histories, multidisciplinary review, and strong public funding. Clinics led by nephrologists performed comparably to multidisciplinary teams when adequately supported. Across all settings, patient selection was more important than the specific type of genetic test used in determining diagnostic success. Kidney genetics services are expanding but remain uneven. This study highlights the need for context-specific, theory-informed, and determinants-targeted strategies to support scalable, equitable, and sustainable genomic care worldwide.

## Introduction

Advancements in genomic science, catalyzed by the Human Genome Project^1^ and accelerated through genomic sequencing technologies,^2^ have revolutionized the landscape of kidney disease diagnostics. These innovations have significantly reduced the cost and turnaround time of genetic testing, enabling broader clinical application. As a result, nephrogenetics has emerged as a distinct subspecialty within nephrology,^3^ with growing evidence supporting the diagnostic, prognostic, and therapeutic utility of genetic testing in both pediatric and adult populations.^4,5^

Despite this momentum, real-world implementation of kidney genetics services remains limited and inconsistent. Integration into routine nephrology care is hindered by unclear models of service delivery, complex diagnostic pathways, and health system-level barriers including workforce limitations, regulatory constraints, and funding challenges.^6–8^ Critically, there is a lack of consolidated evidence on how different models of care are structured, which outcomes they prioritize, and how they are evaluated in terms of service effectiveness, patient-reported outcomes, and implementation success. To address these gaps, we undertook a two-part scoping study comprising:

i. A scoping literature review, using a structured search and synthesis, of published literature on kidney genetics care, and
ii. An international stakeholder consultation of global clinic leads aimed at capturing real-world service structures and implementation experiences and informing gaps in the published literature.

In this scoping study, we synthesize findings from the literature and stakeholder consultation to map the current published landscape; define kidney genetics clinic care models; characterize their geographical and institutional settings; describe reported patient, service, and implementation outcomes; and identify knowledge gaps.

## Methods

### Part 1: Scoping Literature Review

We conducted this review according to a pre-specified protocol, which was prospectively registered on the Open Science Framework (OSF; <u>doi.org/10.17605/OSF.IO/N32VA</u>). The review was conducted and reported following the Joanna Briggs Institute (JBI) methodological guidance for scoping reviews^9^ and the Preferred Reporting items for Systematic reviews and Meta-Analyses extension for Scoping Reviews (PRISMA-ScR) reporting guidelines.^10^ **(Supplemental Table 1)**

#### Eligibility Criteria

We included full-text publications in English, irrespective of study design or geographical location, involving either pediatric or adult populations. Eligible studies were published between January 2000 and 19 February 2025. The decision to include literature from the year 2000 onward was based on the emergence of kidney genetics clinics, growing adoption of genomic sequencing in clinical nephrogenetics during the 2010s and the concurrent development and refinement of key implementation science frameworks in the early 2000s. This timeframe was chosen to ensure the review captured studies reflecting the modern era of genomic testing, contemporary models of kidney genetics care, and implementation framework aligned with current clinical practice, policy, and patient needs.

#### Search Sources and Strategy

A comprehensive search was conducted across three electronic databases: MEDLINE (via PubMed), Scopus, and Embase. The search strategy was developed by R.S.L using a combination of controlled vocabulary and relevant keywords, informed by the Population, Concept, and Context (PCC) framework^9^ and aligned with the review objectives. Two independent medical librarians peer-reviewed the strategy to ensure accuracy and completeness. To identify additional sources, including potentially relevant grey literature, we also conducted a supplementary search using Google Scholar. The complete search strategies for all databases are provided in **Supplemental Table 2**.

#### Study Selection

All retrieved citations were imported into Covidence,^11^ where duplicates were automatically removed. Title and abstract screening were independently performed by four reviewers (R.S.L, J.J, S.J, and A.J.M) to identify potentially eligible studies. Full-text articles were then assessed in detail against the inclusion criteria. Any disagreements were resolved through discussion; unresolved conflicts were adjudicated by a fifth reviewer (E.B), who served as the final decision-maker. Reasons for full-text exclusion were stated.

#### Data Extraction

Two independent reviewers (R.S.L and R.S.Y.L) extracted data independently using a pre-determined data extraction template within Covidence. The extraction form was pilot-tested on a sample of studies and refined iteratively. Extracted data included study characteristics, healthcare context, population, clinic model, genomic testing approach, and reported outcomes. No discrepancies were identified between reviewers during data extraction, and adjudication by a third reviewer was not required. The full list of data extraction fields is provided in **Supplemental Table 3**.

#### Data Analysis and Synthesis

We analyzed and synthesized data across three major thematic areas using a combination of deductive and inductive thematic analysis.^12,13^ First, we mapped the scope, characteristics, and contextual features of included studies using a descriptive summary of key variables. Studies were then grouped into purpose-based thematic domains. Next, we developed a taxonomy of kidney genetics clinic models, informed by predefined structural and functional elements, including team composition, referral pathways, genetic test ordering authority, and mode of service delivery. Classification was guided by decision rules in **Supplemental Table 4**. Finally, we synthesized reported service outcomes, patient-level outcomes, and where available, implementation outcomes. We mapped service and patient-level outcomes to the Institute of Medicine (IOM) framework,^14^ which assesses healthcare quality across six domains: safety, effectiveness, patient-centredness, timeliness, efficiency, and equity. Implementation outcomes were classified using Proctor’s implementation outcomes taxonomy,^15^ which defines constructs essential for evaluating the success of implementation efforts, namely: acceptability, appropriateness, adoption, feasibility, fidelity, cost, penetration, and sustainability. These frameworks were selected to guide consistent outcome categorization across studies.

All coding and thematic synthesis were conducted in Microsoft Excel. A single reviewer performed initial data coding. Frequency counts were performed to quantify the study thematic domains, distribution of clinic models, and reported outcomes.

### Part 2: International Stakeholder Consultation

To complement the scoping review and to inform gaps in the published literature, an international stakeholder consultation with kidney genetics clinic leads, using a structured survey instrument was subsequently undertaken. The consultation was part of the prospectively registered OSF scoping review protocol (as above).

### Stakeholders

Forty-five key kidney genetics clinic leads were identified initially through the literature review of Part 1, professional networks, and existing collaborations. Additional stakeholders were engaged via snowball sampling, whereby existing stakeholders nominated other relevant clinic leads.

#### Survey Development, Structure, and Content

A 42-item survey instrument was developed using Google Forms, informed by the early scoping review findings and expert consultation. The survey captured information across several key domains, including clinic characteristics and structure, patient populations and referral pathways, genetic counseling and testing practices, cascade testing and family management, and service funding models. Stakeholder demographics were also collected; the full questionnaire is provided in in **Supplemental Table 5**.

#### Survey Administration and Data Collection

Survey invitations were distributed via personalized email, with up to three reminder emails sent at monthly intervals to non-respondents. Participation was voluntary. Formal ethics approval was not required as per scoping review methodology. Completion of the survey following the email invitation was considered implied informed consent.

#### Data Analysis

Survey responses were exported into Microsoft Excel for data cleaning and analysis. Descriptive statistics, including frequency counts and proportions, were used to summarize clinic characteristics and service delivery practices.

The complete study protocol and supporting methodological documents, including materials for the international stakeholder consultation, are publicly available via the OSF repository (<u>doi.org/10.17605/OSF.IO/B2VA8</u>)

### Triangulation of Evidence

Findings from the scoping literature review and the international stakeholder consultation were analyzed separately and then triangulated to enhance the completeness and contextual relevance of the evidence. The scoping literature review provided published data, while the stakeholder consultation offered contemporaneous, system-level insights from kidney genetics clinic leads. We compared and synthesized these two data sources thematically, aligning them by clinic models, core service components, and reported outcomes to identify areas of convergence, divergence, and gaps needing further research attention.

## Results

### Existing Literature

#### Study Selection

We identified 4375 studies through systematic searches of MEDLINE (via PubMed), Scopus, Embase, and Google Scholar. After removing 746 duplicates, 3629 titles and abstracts were screened for eligibility. Of these, 148 full-text articles were retrieved and assessed in detail. 88 articles were excluded with reasons documented. Ultimately, 60 studies met the inclusion criteria and were included in this scoping review. The study selection process is illustrated in the PRISMA flow diagram **(Supplemental Figure 1)**.

#### Study Characteristics and Thematic Domains

Among the 60 studies, North America contributed the greatest number (n=23), followed by the Europe (EU) (n=17), Australia/New Zealand (AUS/NZ) (n=10), United Kingdom (UK)/Ireland (n=5), and Asia (n=5) **(Figure 1)**. Supplemental **Figure 2** presents the distribution of publications by year and region, with the earliest study published in 2013. Notably, we saw a steady increase in publications from 2019, reflecting growing interest in kidney genomics research globally.

**Figure 1.**
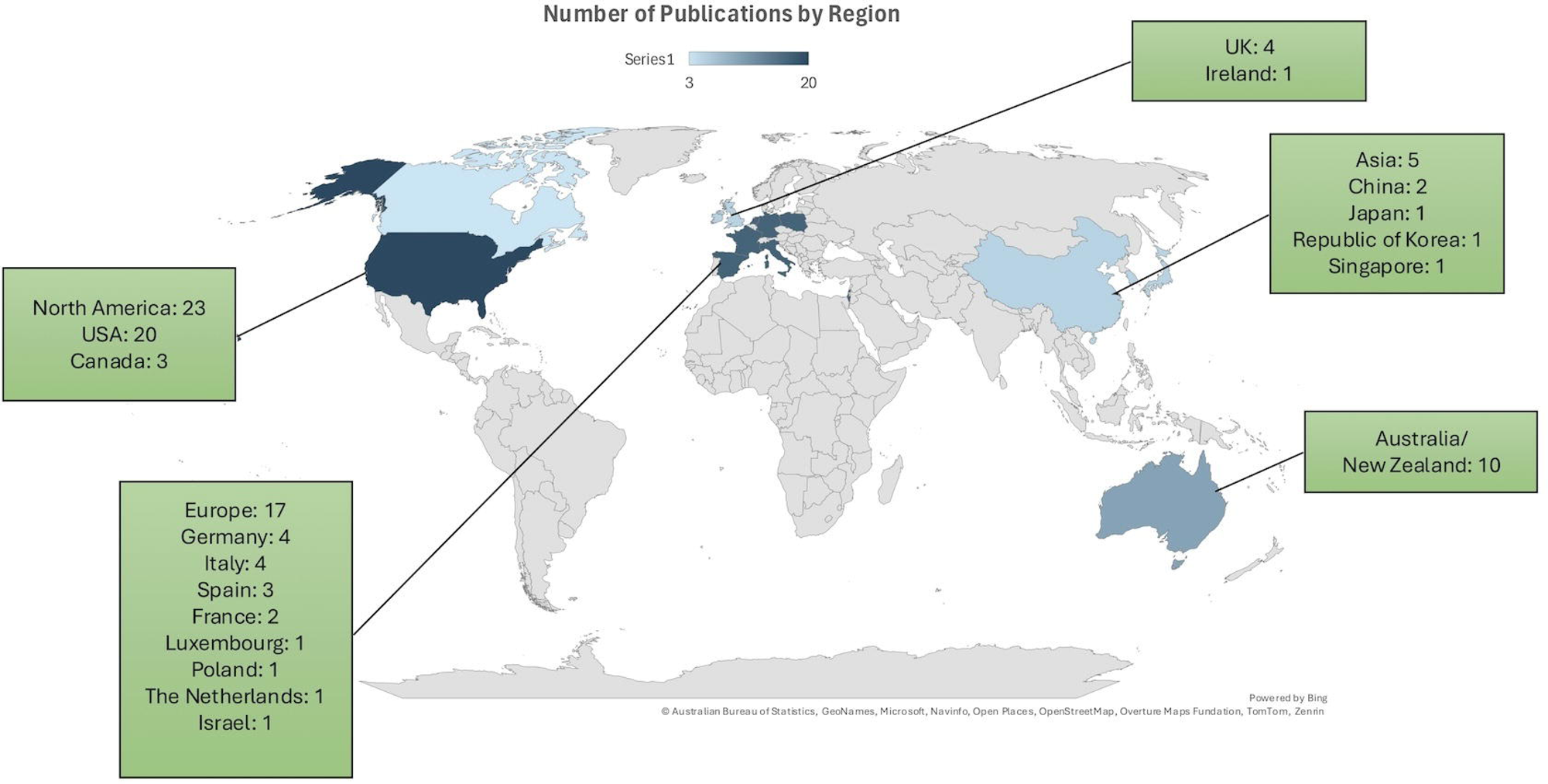
Geographic distribution of included studies by region. This world map illustrates the number of publications (n = 60) included in the scoping review, stratified by region and country. North America contributed the most studies (n = 23; USA = 20, Canada = 3), followed by Europe (n = 17; led by Germany and Italy), Australia/New Zealand (n = 10), the United Kingdom and Ireland (n = 5), and Asia (n = 5; from China, Japan, Republic of Korea, and Singapore). Countries with a single publication included Luxembourg, Poland, The Netherlands, Israel, and Ireland. The shading gradient reflects publication volume per country, with darker colors representing higher numbers. *Note: Israel was grouped under Europe due to its alignment with European research frameworks and professional nephrology networks*.

The studies were grouped into seven main thematic domains **(Supplemental Figure 3)**, with some studies addressing overlapping domains:

i. Kidney genetics clinic model description and evaluation (n=25)
ii. Genomic utility in kidney disease in non-clinic settings (n=26)
iii. Methodology or infrastructure development to support the implementation of kidney genomics (n=33)
iv. Stakeholder experience (n=7)
v. Health economic and system analysis (n=2)
vi. Kidney genomic implementation process evaluation (n=1)
vii. Ethical, legal, and social implications of genetic testing in nephrology (n=1).

**Supplemental Figure 4** shows the grouping of publications by regions, first authors, and thematic domains. Detailed study characteristics across each domain are summarized in **Table 1**, **Supplemental Tables 6-9.**

**Table 1:**
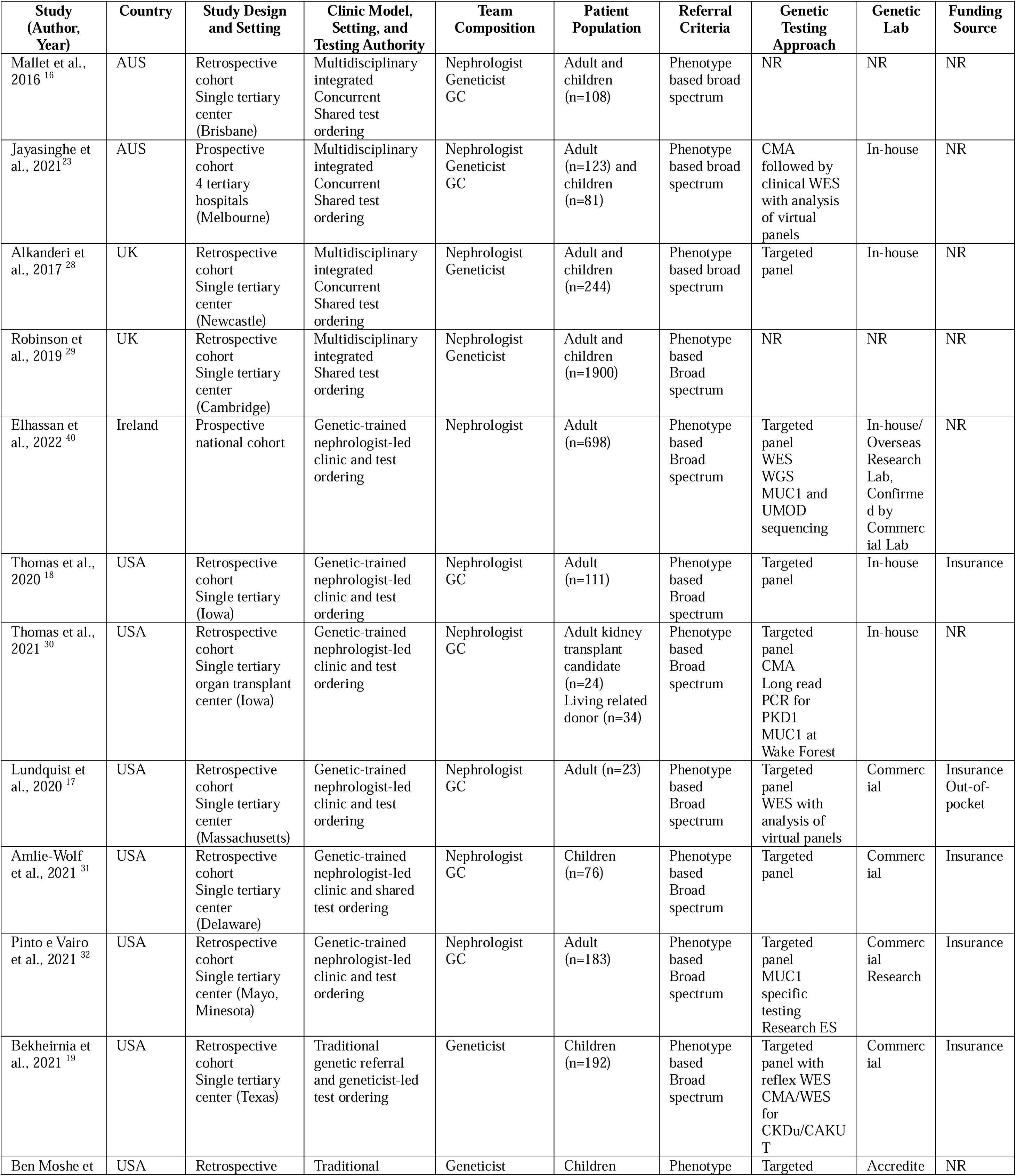
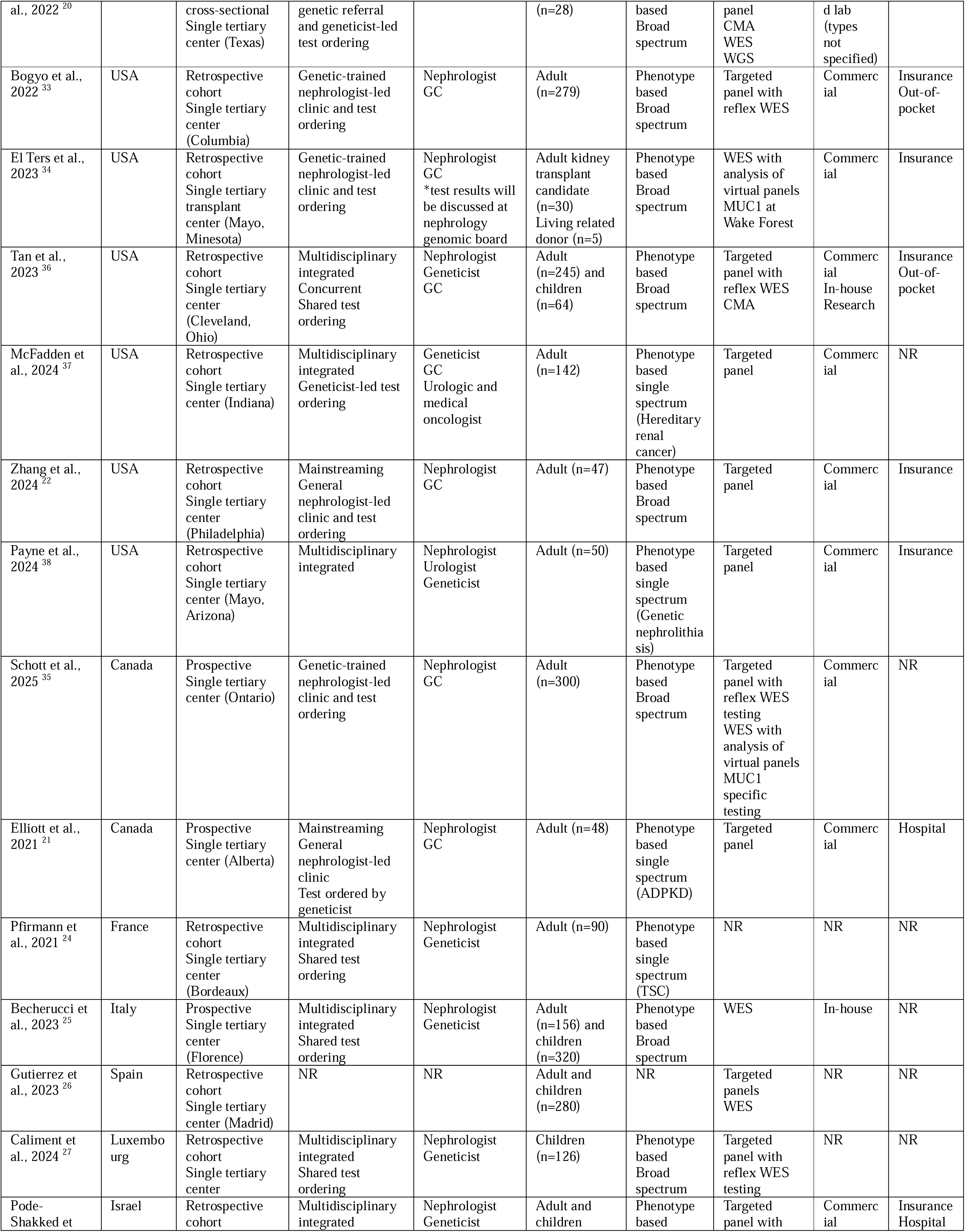
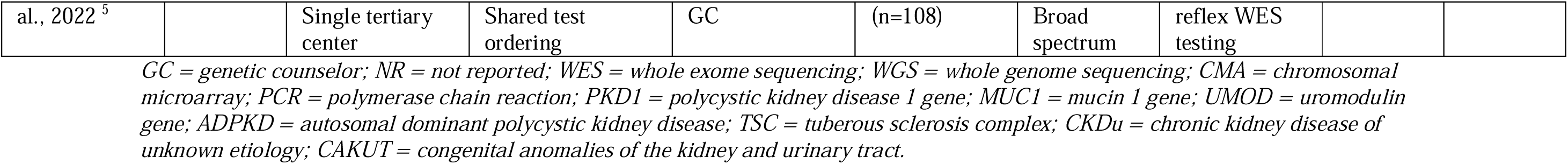
Characteristics of studies describing and evaluating kidney genetics clinic models (n=25 studies)

### Survey respondents

We analyzed 48 completed surveys: 43 respondents from the initial survey invitation (43/45 invited; 96% response rate) and 5 via snowball sampling. By region, responses were most frequent from AUS/NZ 16/48 (33%), followed by the UK/Ireland 12/48 (25%), North America 11/48 (23%), EU 7/48 (15%), and Asia 2/48 (4%) **(Supplemental Figure 5a)**. Professional roles were predominantly adult nephrologist 26/48 (54%), clinical geneticist 8/48 (17%), pediatric nephrologist 6/48 (13%), nephrologist-geneticist 6/48 (13%), genetic counselor 1/48 (2%), and laboratory geneticist 1/48 (2%). Nearly half reported 10 years or more of experience in nephrogenetics 21/48 (44%), 7-9 years in 17/48 (35%), 4-6 years in 7/48 (15%), and 1-3 years in 3/48 (6%) **(Supplemental Figure 5b)**.

### Clinic Models

#### Taxonomy

Using our predefined taxonomy **(Supplemental Table 4)**, four clinic models were identified across the 25 studies **(Table 1)**: (i) multidisciplinary integrated clinics; (ii) nephrologist-led clinics (genetics-trained nephrologists with genetic team support); (iii) mainstreaming within general nephrology; and (iv) traditional genetics referral models. In the literature, multidisciplinary clinics were the first reported in 2016^16^ and remain the most frequently published (12 studies), followed by nephrologist-led clinics, which first appeared in 2020^17,18^ (9 studies). Traditional genetics referral and mainstreaming models each have two reports since 2021^19–22^ **(Figure 2)**. Survey data show a similar ranking: multidisciplinary clinics were most common among respondents (n=24), followed by nephrologist-led (n=15), mainstreaming (n=4), and hybrid model (n=2), the latter refers to services that combine elements of multidisciplinary, nephrologist-led, and/or mainstreaming approaches. **(Figure 3a)**. Taken together, multidisciplinary clinics predominate globally, with nephrologist-led clinics growing rapidly; mainstreaming is emerging but remains less common.

**Figure 2.**
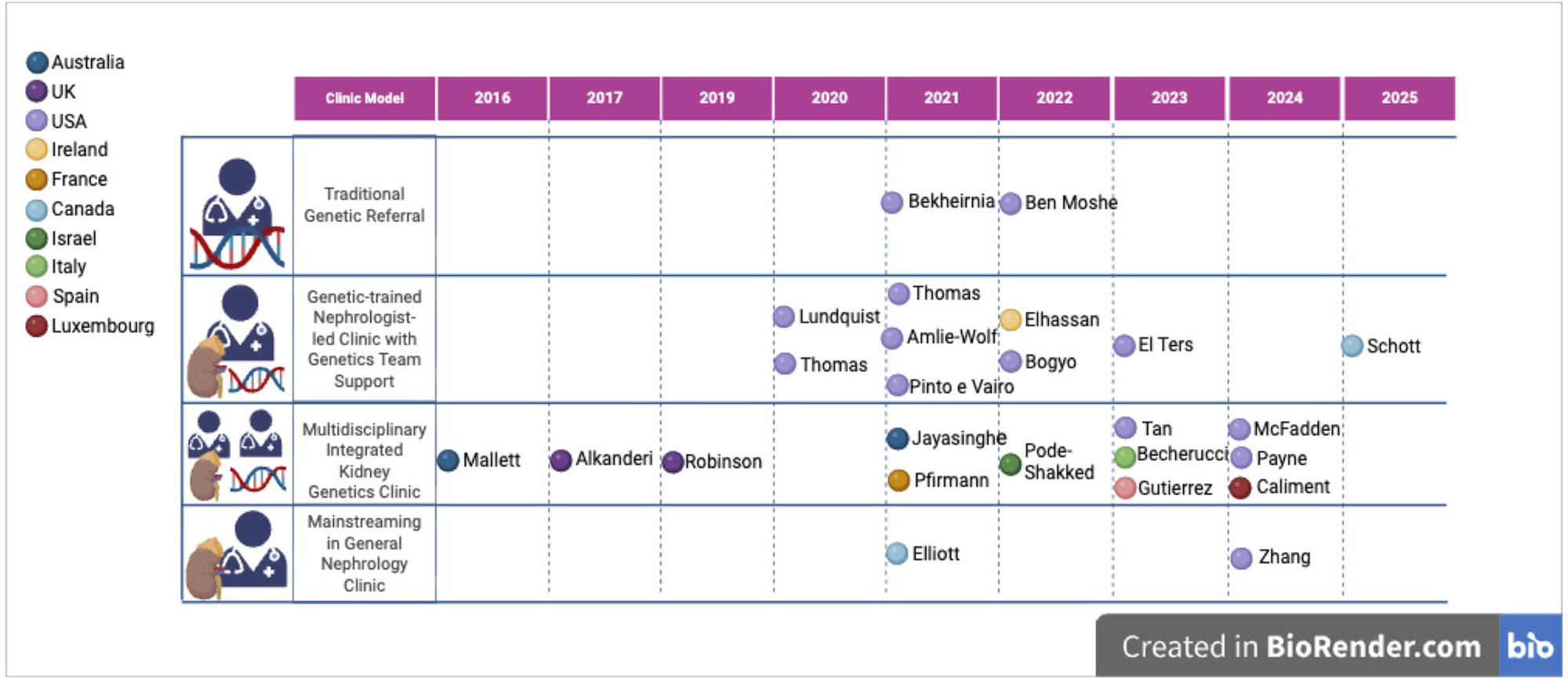
Temporal trend of published kidney genetics clinic models by type and country (2016–2025) Timeline of 25 peer-reviewed publications describing or evaluating kidney genetics clinics, categorized by clinic model and year of publication. Clinic models are represented in rows: traditional genetic referral, genetic-trained nephrologist-led clinic with genetics team support, multidisciplinary integrated kidney genetics clinic, and mainstreaming in general nephrology clinic. Country of origin is indicated by color-coded dots: Australia (dark blue), UK (purple), USA (lavender), Ireland (gold), France (orange), Canada (light blue), Israel (green), Italy (olive), Spain (pink), and Luxembourg (maroon). The earliest publication was from Australia in 2016 (Mallett et al.), with a growing diversity of models and geographic representation over time.

**Figure 3a.**
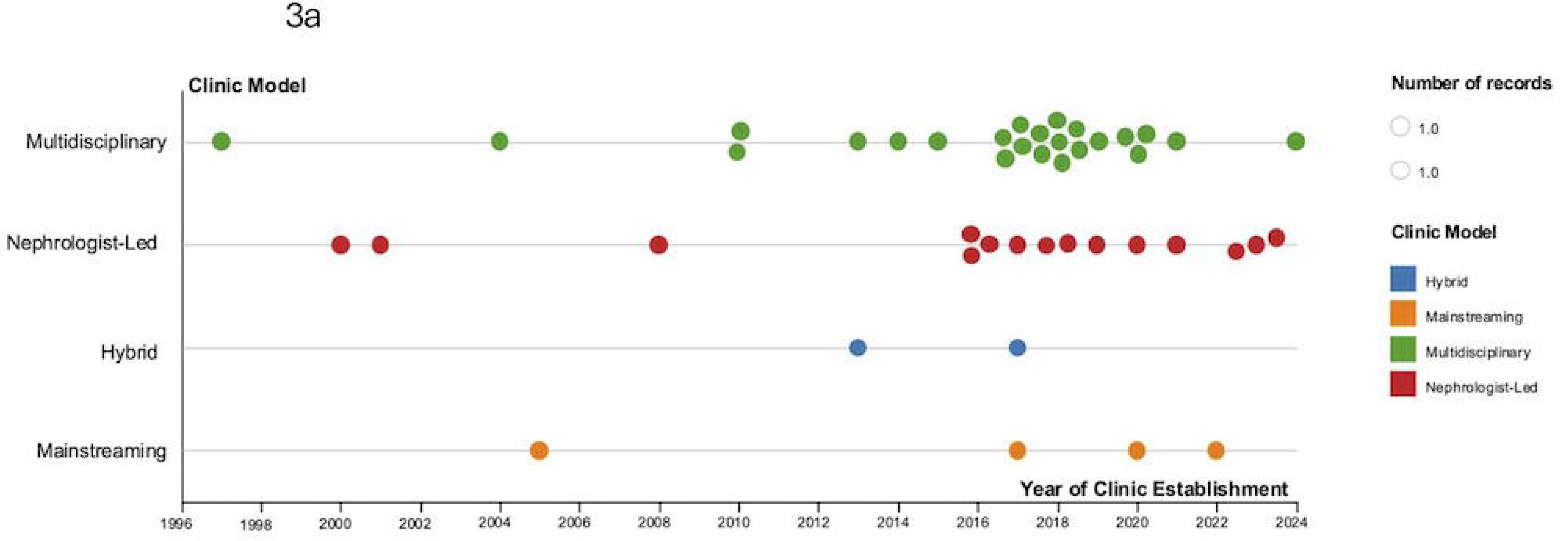
Timeline of kidney genetics clinic establishment by clinic model (survey respondents, n = 45; missing response, n=3) Each dot represents a unique clinic, plotted by year of establishment and categorized by clinic model: multidisciplinary (green), nephrologist-led (red), hybrid (blue), and mainstreaming in general nephrology (orange). Multidisciplinary clinics were the earliest established, dating back to 1997, while nephrologist-led models have increased in number since 2015. Fewer hybrid and mainstreaming clinics were reported. Hybrid model refers to services that combine elements of multidisciplinary, nephrologist-led, and/or mainstreaming approaches.

#### Temporal Trends

Survey timelines indicate the earliest clinics in the UK/Ireland in 1997, followed by EU in 2001, with subsequent expansion in AUS/NZ from 2013 onward, North America from 2017, and more recent growth in Asia since 2020 **(Figure 3b)**. By model **(Figure 3a)**, multidisciplinary clinics span the entire period and cluster most densely from 2016 onward; nephrologist-led services appear after 2016 with steady uptake; mainstreaming entries are recent and sparse; hybrid models are infrequent across the last decade.

**Figure 3b.**
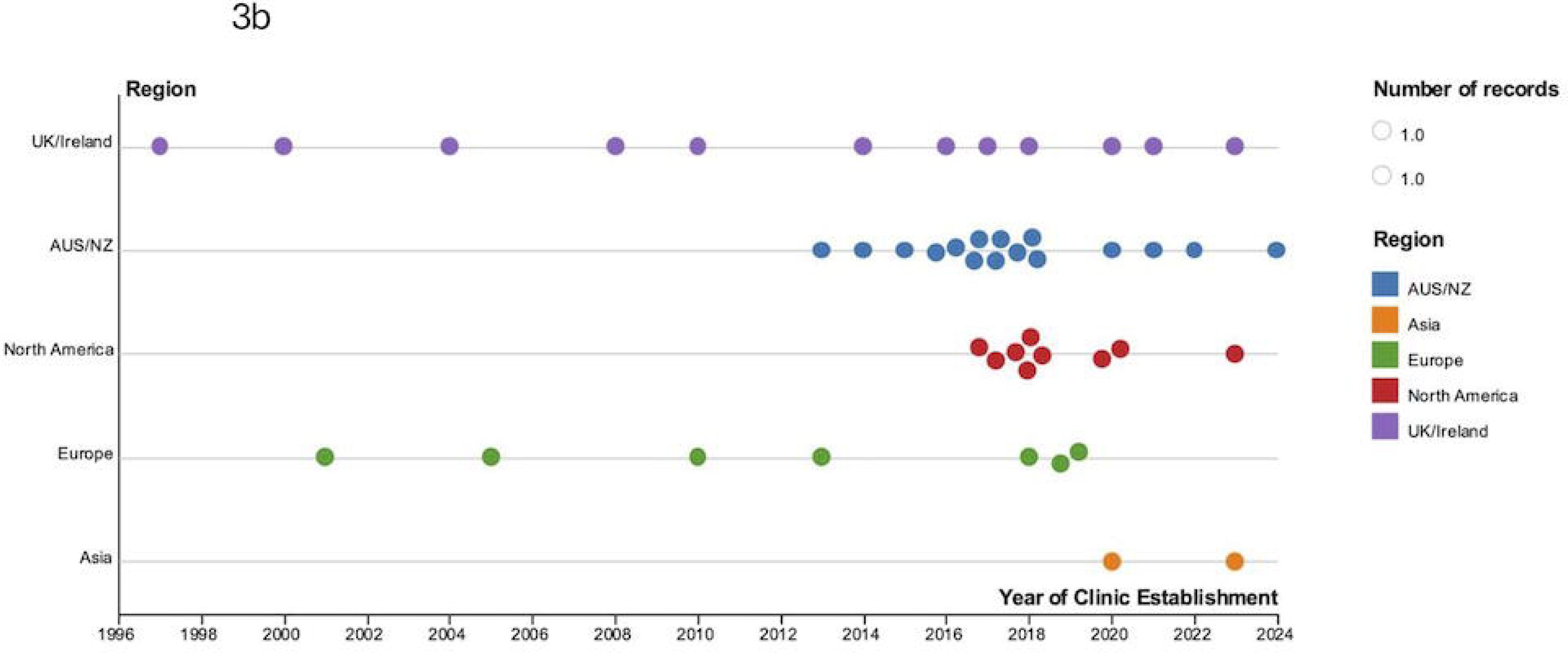
Timeline of kidney genetics clinic establishment by region (survey respondents, n = 46; missing response = 2) Each dot represents a clinic, plotted by year of establishment and stratified by region: UK/Ireland (purple), Australia/New Zealand (blue), North America (red), Europe (green), and Asia (orange). UK/Ireland and Europe reported the earliest established clinics, with rapid growth across all regions beginning in the late 2010s. Regional differences in timing reflect varying stages of service development and implementation.

#### Regional Distribution

When stratified by region, publications from AUS/NZ and EU described multidisciplinary clinics^5,16,23–27^ exclusively (100%), and two-thirds of UK/Ireland reports were multidisciplinary.^28,29^ In North America, just over half of publications were nephrologist-led (54%),^17,18,30–35^ with the remainder multidisciplinary (20%), ^36–38^ mainstreaming (13%),^21,22^ or traditional genetics referral models (13%)^19,20^ **(Figure 4a)**. In contrast, the survey suggests a more diversified contemporary landscape: AUS/NZ remains predominantly multidisciplinary (75%), with smaller proportions nephrologist-led and mainstreaming (12.5% each); North America and UK/Ireland show an approximately even split between multidisciplinary and nephrologist-led models; EU also shows both models; and the two respondents from Asia reported nephrologist-led clinics **(Figure 4b)**. Overall, published reports emphasize early adoption of multidisciplinary clinics, whereas current practice appears more diversified with signals of increasing nephrologist-led services across several regions.

**Figure 4a.**
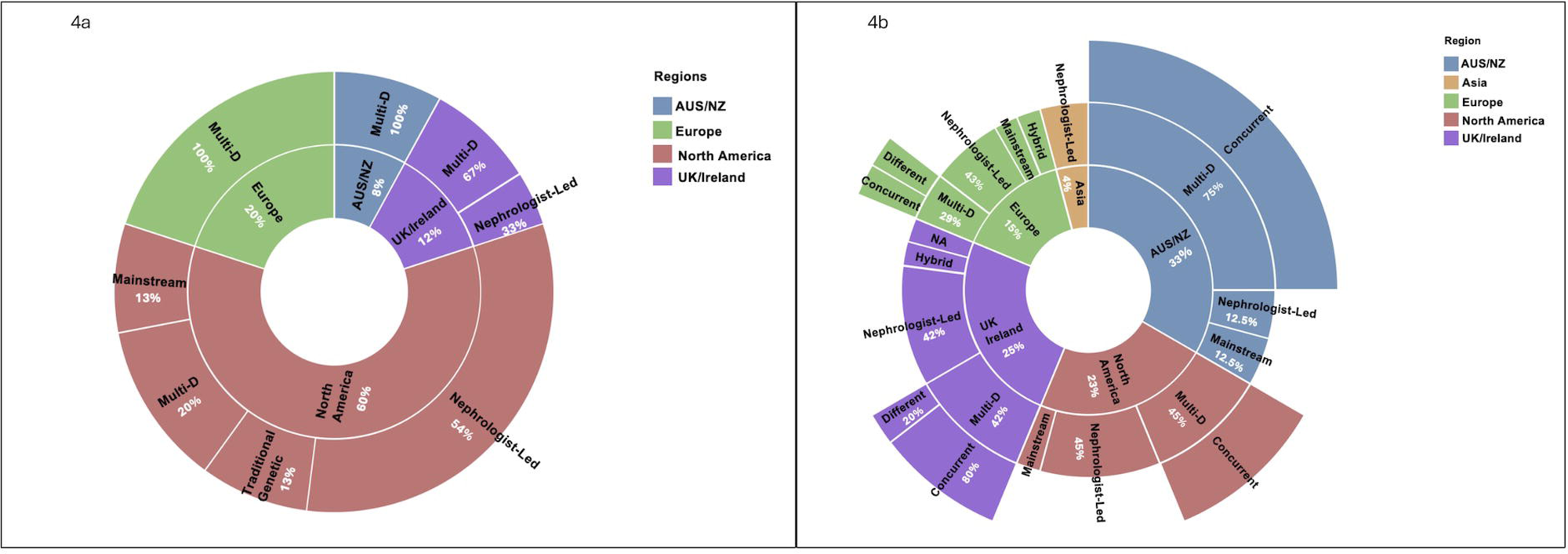
Regional distribution of kidney genetics clinic models reported in the literature (n = 25) Sunburst chart illustrating the proportion of published kidney genetics clinic models by region and clinic type. Color coding represents regions: Australia/New Zealand (AUS/NZ, blue), Europe (green), UK/Ireland (purple), and North America (red). Clinic models include multidisciplinary (Multi-D), nephrologist-led, traditional genetic referral, and mainstreaming. Most European and AUS/NZ clinics were multidisciplinary (100%), while North America showed the greatest diversity, with a predominance of nephrologist-led clinics (54%), followed by Multi-D (20%), traditional genetic referral (13%), and mainstreaming (13%). UK/Ireland clinics included both Multi-D and nephrologist-led models.

**Figure 4b.**
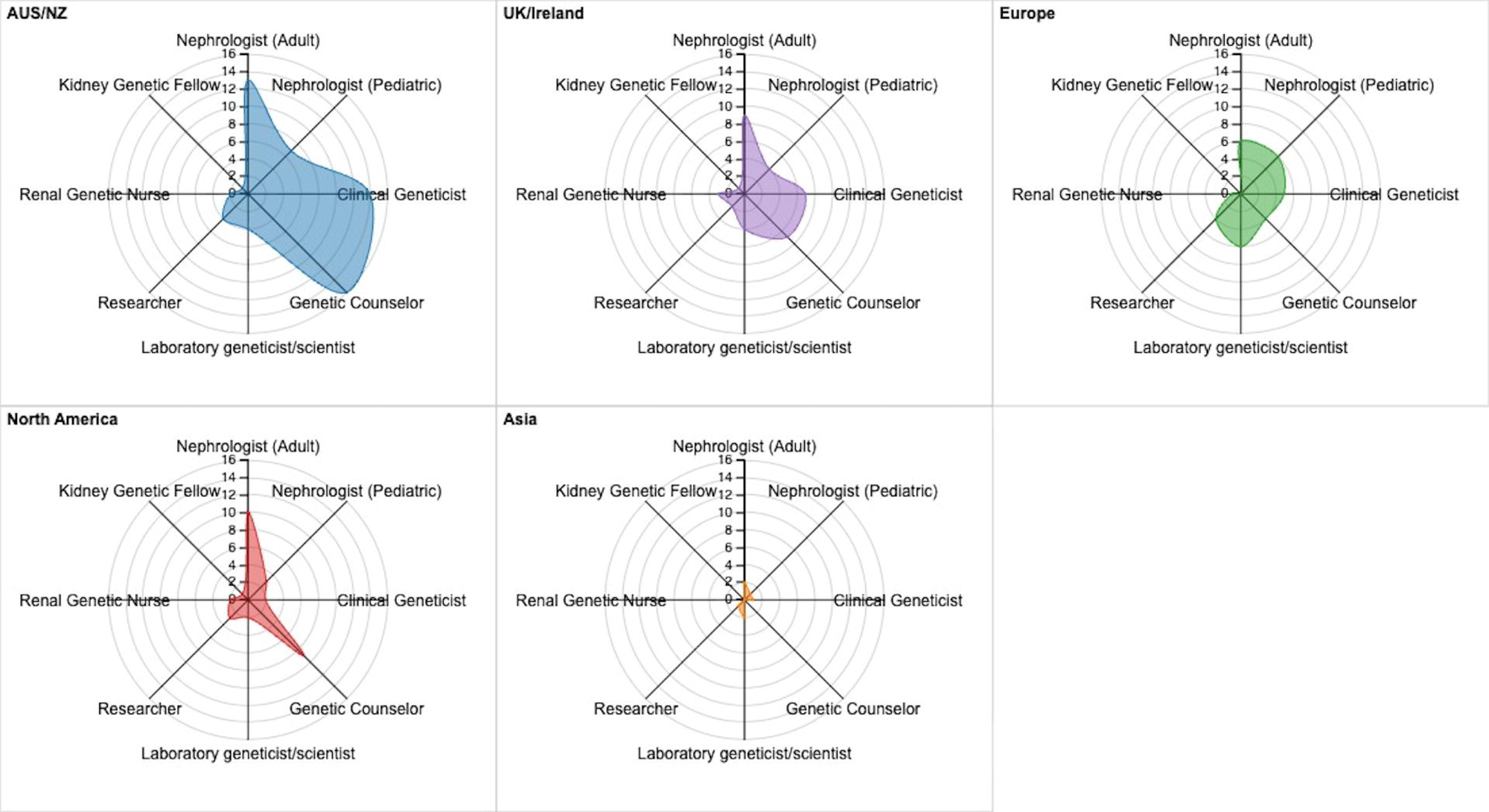
Regional distribution of kidney genetics clinic models and clinic setting reported by global clinic leads (survey respondents, n = 47; missing response, n = 1) Sunburst chart displaying the clinic models and operational settings by region based on survey responses. The innermost ring indicates region: Australia/New Zealand (AUS/NZ, blue), Asia (orange), Europe (green), North America (red), and UK/Ireland (purple). The middle ring shows the clinic model: multidisciplinary (Multi-D), nephrologist-led, mainstream, or hybrid. Hybrid model refers to services that combine elements of multidisciplinary, nephrologist-led, and/or mainstreaming approaches. The outermost ring indicates the setting of the multidisciplinary clinic, i.e., whether multidisciplinary providers conduct the clinic concurrently (on the same day), on different days, or not applicable (NA) for non-multidisciplinary models. AUS/NZ clinics were mostly multidisciplinary with concurrent setups, while North America showed a mix of models and settings. UK/Ireland and Europe exhibited varied clinic structures and configurations.

#### Team Composition

Across the scoping review **(Table 1)** and survey **(Figure 5)**, services in AUS/NZ, the UK/Ireland, and EU were typically multidisciplinary—bringing together adult and pediatric nephrologists, clinical geneticists, and genetic counselors, with renal genetic nurses/trainees and laboratory liaisons variably included. In North America, nephrologist-led clinics were more common, usually supported by genetic counselors with less routine involvement of clinical geneticists. In Asia, respondents frequently described resource-constrained teams in which a genetics-trained nephrologist leads the service and often undertakes genomic evaluation and counselling due to limited access to clinical geneticists and genetic counselors.

**Figure 5.**
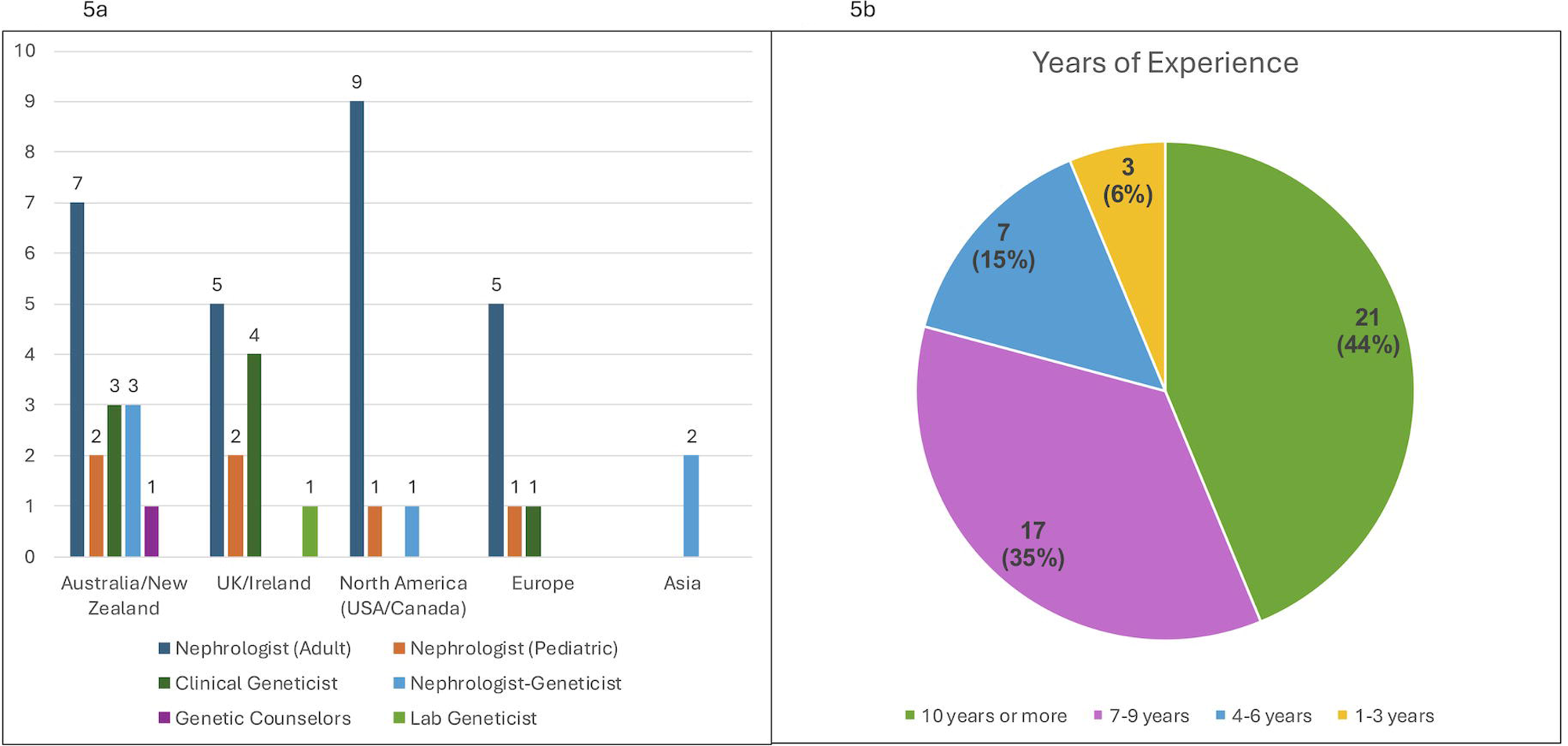
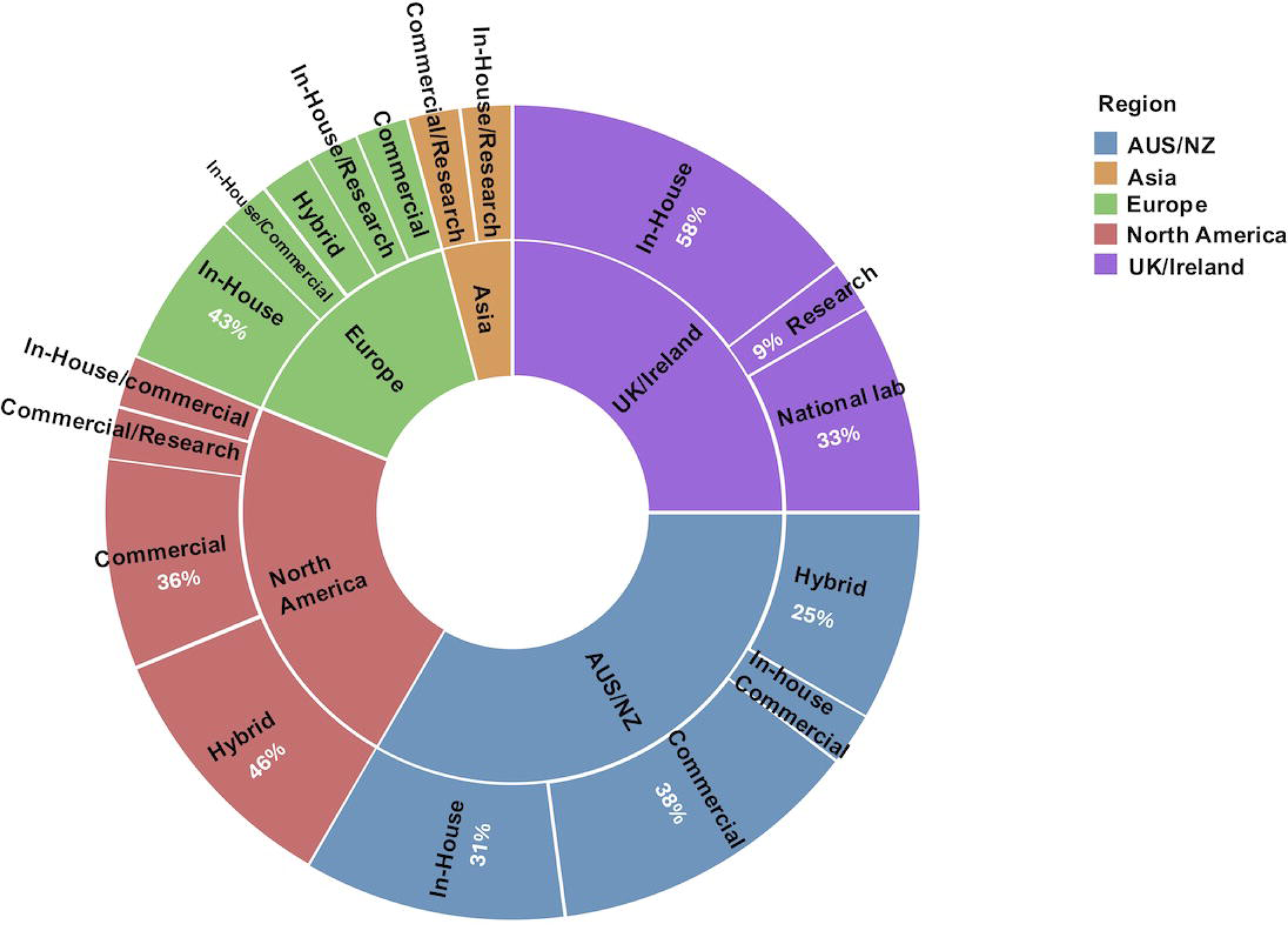
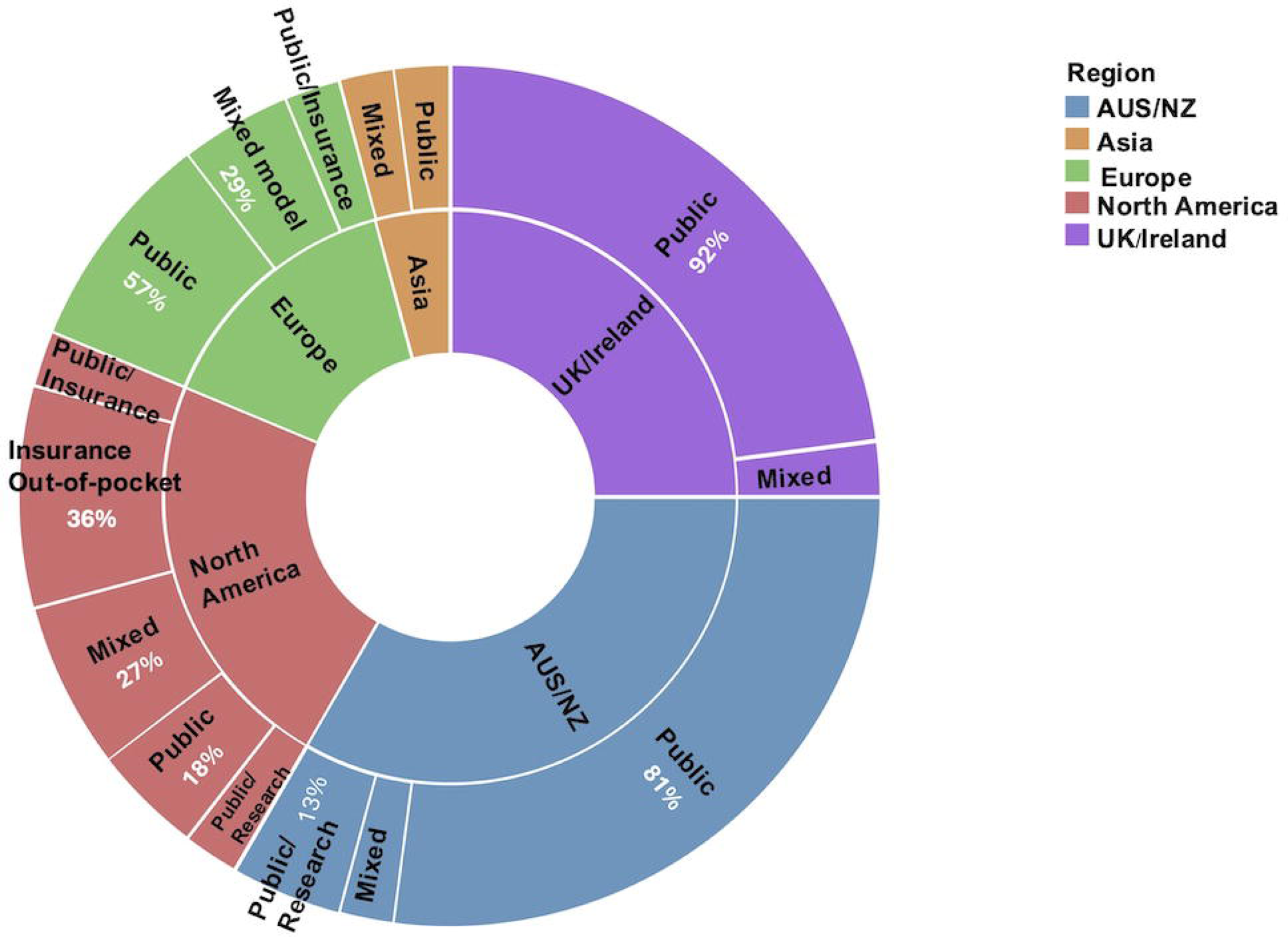
Team composition in kidney genetics clinics, stratified by region (survey respondents, n = 48) Radar plots illustrate the frequency of multidisciplinary team members involved in kidney genetics clinics across five regions: Australia/New Zealand (AUS/NZ, blue), Asia (orange), Europe (green), North America (red), and UK/Ireland (purple). Reported roles include adult and pediatric nephrologists, kidney genetics fellows/trainees, renal genetic nurses, clinical geneticists, genetic counselors, laboratory geneticists/scientists, and researchers. AUS/NZ, UK/Ireland, and Europe were more likely to report multidisciplinary team composition including clinical geneticists and genetic counselors. In contrast, North American clinics were more commonly led by nephrologists with genetic counselor support and less frequent involvement of clinical geneticists. Asia reported limited workforce representation, with most clinics anchored by nephrologists.

#### Testing Approach

Genetic testing strategies were heterogeneous across studies, reflecting resource constraints and local practice **(Table 1)**. In our international stakeholder consultation, comprehensive approaches using whole exome sequencing (WES)/whole genome sequencing (WGS) with virtual panels were common in AUS/NZ and UK/Ireland (8/16 [50%] and 6/11 [55%] of respondents with data, respectively) and frequent in EU (3/7 [43%]) **(Figure 6)**. By contrast, North American clinics more often reported phenotype-guided targeted panels, with optional reflex to WES/WGS (6/11 [55%]). Hybrid testing approaches–combining testing modalities such as WES, WGS, and targeted panels–were used by a minority across regions (range 9–36%; AUS/NZ 3/16, UK/Ireland 1/11, EU 2/7, North America 4/11), and research testing with reflex clinical confirmation was uncommon (observed in 1/11 [9%] North American site). In Asia (n=2), one clinic reported comprehensive WES/WGS with virtual panels and one used targeted panels.

**Figure 6.**
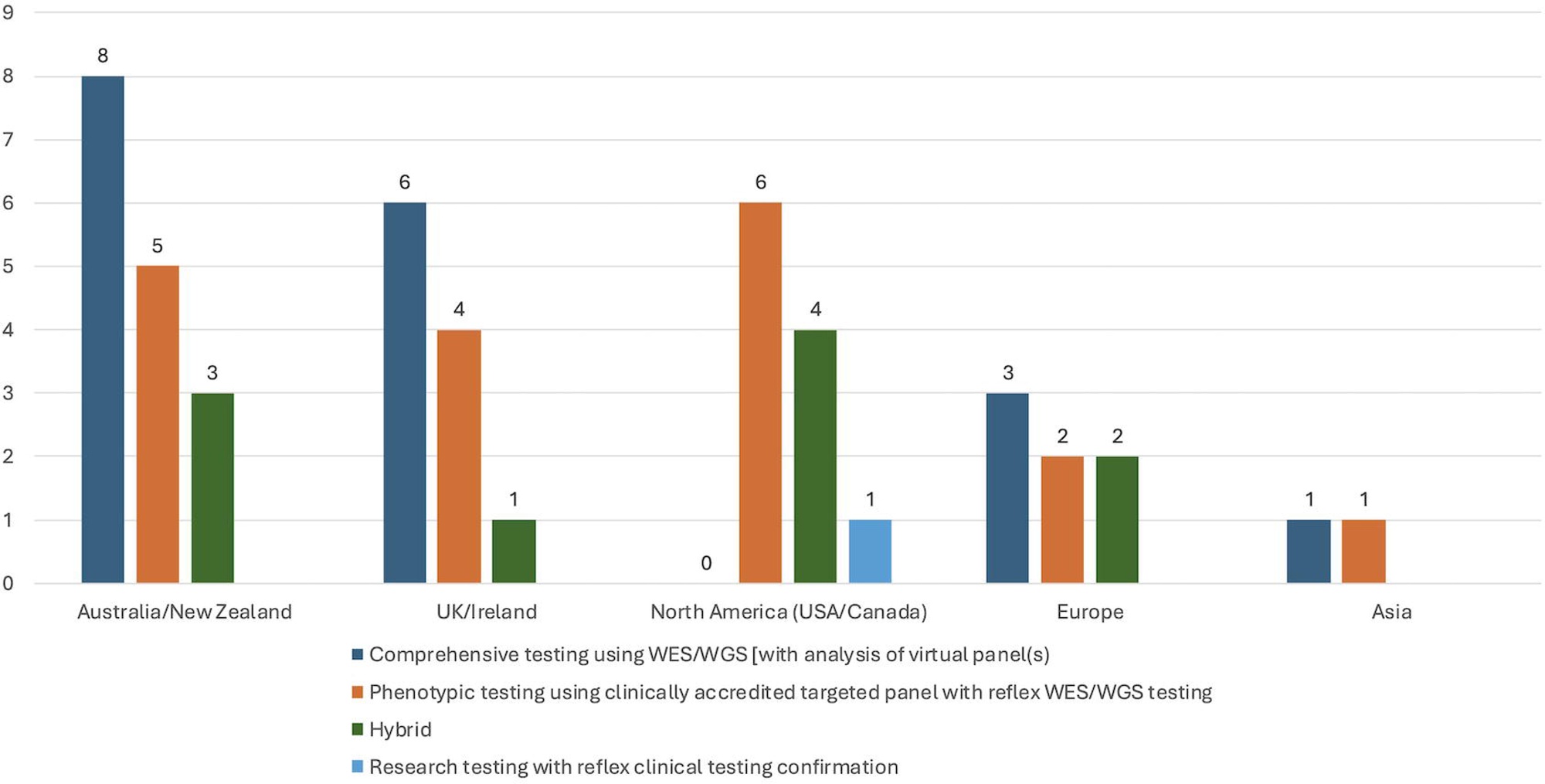
Genetic testing approaches used in kidney genetics clinics, stratified by region (survey respondents, n = 47; missing response, n=1) Bar chart showing the distribution of genetic testing approaches reported by clinics across five regions. Testing strategies were classified as follows: comprehensive testing using whole exome sequencing (WES) or whole genome sequencing (WGS) with virtual panel analysis (dark blue), phenotypic testing using clinically accredited targeted panels with reflex to WES/WGS (orange), hybrid testing approach–combining testing modalities such as WES, WGS, and targeted panels (green), and research testing with reflex clinical confirmation (light blue). Australia/New Zealand, UK/Ireland, and Europe most frequently reported comprehensive testing, while North America predominantly used phenotypic panel-based testing. Asia showed limited testing strategies.

#### Genetic Laboratory

Laboratory providers were variably reported in the literature; where stated, North American studies most commonly used commercial laboratories (12/15, 80%; Table 1).^17,19,21,22,31–33,35–39^ Survey data showed regional contrasts **(Supplemental Figure 6)**. UK/Ireland clinics predominantly used public providers—in-house hospital or national laboratories (91% of respondents)—with limited use of research-only labs. Europe was mixed, with an in-house plurality (43%). In AUS/NZ, use was split across in-house (31%), commercial (38%), and hybrid models (in-house/commercial ± research, 25%). In North America, a commercial plurality was observed (36%), alongside substantial hybrid use (46%). Asian responses (n=2) reflected hybrid arrangements (in house/commercial/research).

#### Funding

Reporting of test funding sources was limited in the literature; where stated, North American studies most often cited private payers (health insurance or patient out-of-pocket).^17–19,22,31–34,36,38^ Survey data showed a contrasting pattern across regions **(Supplemental Figure 7)**. Public funding predominated in the UK/Ireland (92%) and was common in AUS/NZ (81%) and EU (57%). In North America, private payment (insurance/out-of-pocket) was the single most frequently reported category (36%), with the remainder split between mixed models (public ± private; 27%) and publicly supported pathways (including public and research). Asian responses (n=2) indicated a split between public and mixed funding (each 50%).

### Outcomes

Across 25 studies, outcome reporting was dominated by diagnostic yield: 23/25 (92%) reported yield.^5,16–19,21–28,30–38,40^ Timeliness measures were less common—4/25 (16%) reported wait time from referral to clinic review,^23,31,33,35^ and 8/25 (32%) reported time from genetic assessment to receipt of a genetic diagnosis.^17,21,23,26,27,31,35,40^ Patient-reported outcomes were rarely evaluated (overall clinic satisfaction in 3/25 [12%]).^21,24,29^ Implementation outcomes were similarly rarely reported, except for test penetration (see below); only 1/25 (4%) assessed fidelity (adherence to recommended end-organ surveillance timelines).^24^ Details are summarised in **Table 2**.

**Table 2:** Reported outcomes across studies evaluating kidney genetics clinic models (n=25 studies)

| Study (Author, Year) | Country | Number of Patients Offered Test | Number of Patients Sequenced | Number of Patients with Genetic Diagnosis | Diagnostic Yield (%) | Test Penetration Rate (%) | Turn-Around-Time | Patient Satisfaction | Implementation Outcomes other than Penetration |
| --- | --- | --- | --- | --- | --- | --- | --- | --- | --- |
| Mallet et al., 2016<sup>16</sup> | AUS | 108 | 75 (83 tests) | 32 | 39 | 69 | NR | NR | NR |
| Jayasinghe et al., 2021<sup>23</sup> | AUS | 225 | 204 | 80 | 39 | 91 | Median time from referral to clinic review 2.4 months (0-16.9)<br>Median time from WES request to result report 6 months (10 days-15.8 months) | NR | NR |
| Alkanderi et al., 2017<sup>28</sup> | UK | 62 | 62 | 26 | 42 | 100 | NR | NR | NR |
| Robinson et al., 2019<sup>29</sup> | UK | 1900 | NR | NR | NR | NR | NR | Staff 97% (n=170)<br>Service 97.7% (n=145) | NR |
| Elhassan et al., 2022<sup>40</sup> | Ireland | 698 | 677 | 348 | 51.4 | 97 | Median time from clinic review to return of results 131 days (IQR 100-205) | NR | NR |
| Thomas et al., 2020<sup>18</sup> | USA | 58 | 43 | 26 | 60 | 74 | NR | NR | NR |
| Thomas et al., 2021<sup>30</sup> | USA | 24 kidney transplant recipients<br>34 living-related donors | 24 kidney transplant candidates<br>20 living-related donors | 12 transplant candidates<br>7 living-related donors | 50 (transplant candidates)<br>30.5 (living-related donors) | 100 (kidney transplant recipients)<br>59% (living-related donors) | NR | NR | NR |
| Lundquist et al., 2020<sup>17</sup> | USA | 23 | 19 | 9 | 47 | 83 | Mean time from sample submission to result return 55 days (9-174)<br>Average time to coordinate insurance coverage or out-of-pocket payment before initiation of testing: 17 days | NR | NR |
| Amlie-Wolf et al., 2021<sup>31</sup> | USA | 102 | 76 (86 tests) | 28 | 33 | 75 | Median time from referral to sample collection 15 days (IQR 4-36)<br>Median time from sample collection to results 22 days (IQR 15-39)<br>Median time from request to results 40 days (IQR 27.7-62) | NR | NR |
| Pinto e Vairo et al., 2021<sup>32</sup> | USA | 183 | 163 | 50 | 30.7 | 89 | NR | NR | NR |
| Bekheirnia et al., 2021<sup>19</sup> | USA | 187 | 158 | 81 | 51 | 84.5 | NR | NR | NR |
| Ben Moshe et al., 2022<sup>20</sup> | USA | NR | 28 | NR | NR | NR | NR | NR | NR |
| Bogyo et al., 2022<sup>33</sup> | USA | 231 | 210 | 57 | 27 | 91 | Average wait time from referral to clinic review: 37 days | NR | NR |
| El Ters et al., 2023<sup>34</sup> | USA | NR | 30 | 13 | 43.3 | NR | NR | NR | NR |
| Tan et al., 2023<sup>36</sup> | USA | 309 | 295<br>256 had result available | 115 | 45 | 95.4 | NR | NR | NR |
| McFadden et al., 2024<sup>37</sup> | USA | 142 | 133 | 49 | 36.8 | 94 | NR | NR | NR |
| Zhang et al., 2024<sup>32</sup> | USA | NR | 47 | 24 | 51 | NR | NR | NR | NR |
| Payne et al., 2024<sup>38</sup> | USA | 50 | 33 | 9 | 27 | 66 | NR | NR | NR |
| Schott et al., 2025<sup>35</sup> | Canada | 322 | 300 | 103 | 34.3 | 93 | Median wait time from referral to clinic review: 4.1 months (IQR: 5.2)<br>Median time from genetic assessment to result: 2.9 months (IQR: 6.3) | NR | NR |
| Elliott et al., 2021<sup>21</sup> | Canada | 26 | 18 | 11 | 61 | 69 | Median time from referral to return results: 184 days | Service 100% (n=11) | NR |
| Pfirmann et al., 2021<sup>24</sup> | France | NR | 32 | 23 | 72 | NR | NR | Service 100% (n=12) | Fidelity of target organ evaluation according to recommendation: 32-100% |
| Becherucci et al., 2023<sup>50</sup> | Italy | 519 | 476 | 319 | 67 | 92 | NR | NR | NR |
| Gutierrez et al., 2023<sup>26</sup> | Spain | 280 | 86 | NR | 25.7 | 31 | Mean time from test request to result: 2 months | NR | NR |
| Caliment et al., 2024<sup>27</sup> | Luxembourg | 93 | 93<br>44 had result available | 28 | 63 | 100 | Average time to obtain genetic test results was 6 months | NR | NR |
| Pode-Shakked et al., 2022<sup>5</sup> | Israel | 108 | 107<br>74 had results available | 42 | 57 | 99 | NR | NR | NR |

NR = not reported; WES = whole exome sequencing; WGS = whole genome sequencing; CMA = chromosomal microarray; IQR = interquartile range.
Diagnostic yield (%) refers to the proportion of patients sequenced who received a genetic diagnosis. Test penetration rate (%) refers to the proportion of patients offered testing who underwent sequencing. Turn-around-time represents the reported duration from referral, sample collection, or test request to result return; reporting format and starting points vary across studies. Patient satisfaction reflects staff or service-level satisfaction metrics. Implementation outcomes refer to measures such as fidelity to care recommendations, where assessed.
Note: Due to variability in reporting methods and clinical workflows, the time points used to calculate turn-around-time (e.g., referral-to-review, sample-to-result, or test request-to-result) are not uniform across studies.

### Diagnostic Yield and Test Penetration

Beyond yield, we examined test penetration—defined as the proportion of eligible patients offered testing who completed testing. Twenty of 25 studies reported both outcomes and were included in comparative analyses.^5,16–19,21,23,25–28,30–33,35–38,40^ We summarised yield and test penetration overall and stratified by region, clinic model, and testing approach in the following sections. Definitions of “eligibility” and offer pathways varied across studies; where possible, we used authors’ definitions and extracted study-level numerators and denominators (details in **Table 2**).

#### Yield vs Test Penetration by Region

Test penetration was generally high across regions, whereas diagnostic yield was more variable **(Figure 7)**. North American studies clustered at moderate yields (27–61%) with consistently high penetration (66-100%).^17–19,21,30–33,35–38^ European studies tended to report higher yields (57-67%) with very high penetration (92-100%),^5,25,27^ aside from one outlier with lower uptake (31%) at a modest yield (26%).^26^ UK/Ireland (n=2) showed high penetration (97-100%) at intermediate yields (42-51%).^28,40^ AUS/NZ (n=2) reported intermediate yields (39%) with mid-to-high penetration (69-91%).^16,23^

**Figure 7.**
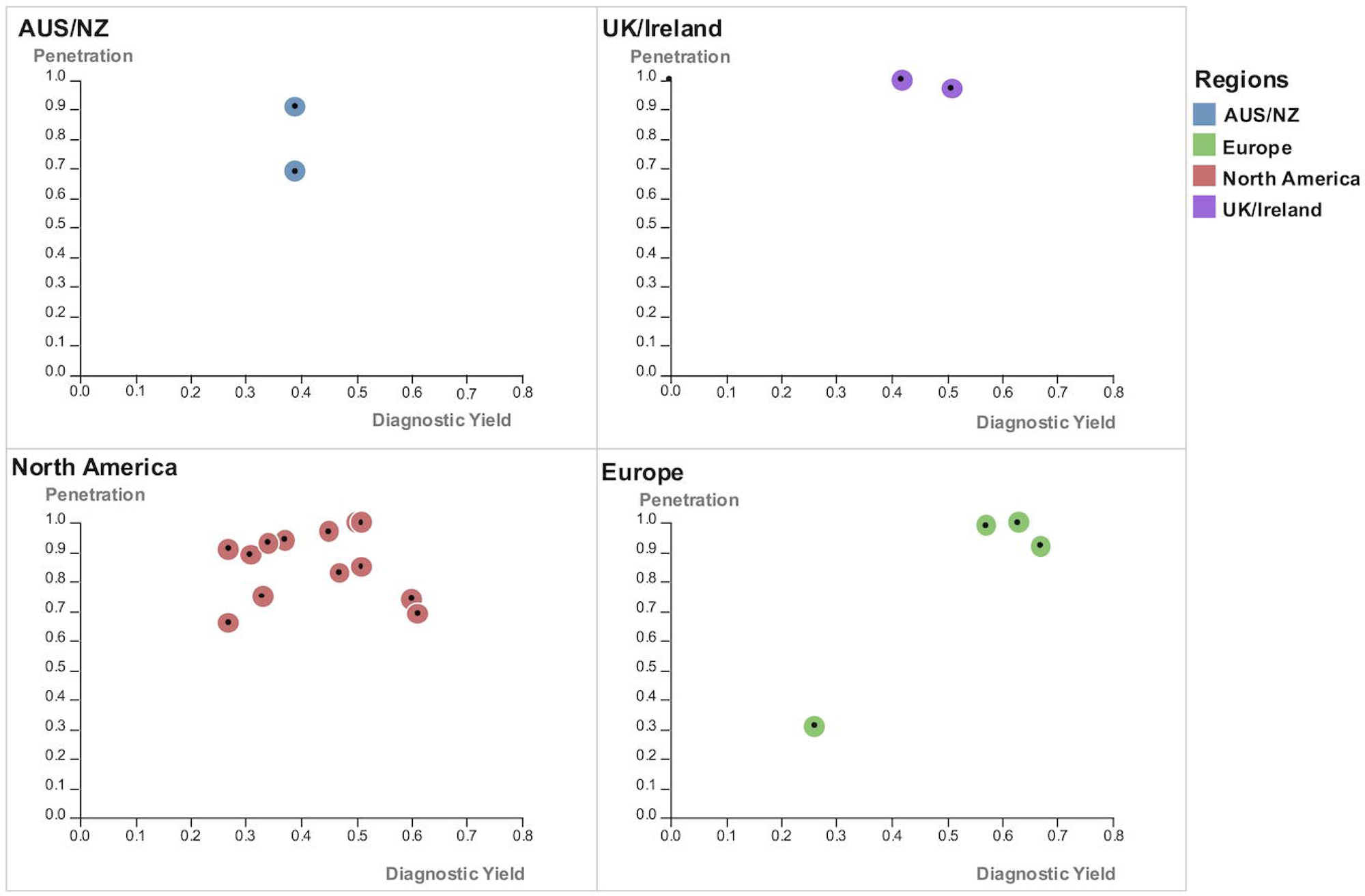
Diagnostic yield versus test penetration in kidney genetics clinics, stratified by region (literature review, n = 20 studies) Scatter plots display diagnostic yield (x-axis) against genetic test penetration rate (y-axis) for 20 studies reporting both outcomes. Each dot represents one study, color-coded by region: Australia/New Zealand (AUS/NZ, blue), UK/Ireland (purple), North America (red), and Europe (green). Studies from Europe generally demonstrated high diagnostic yield and high penetration rates, with consistent clustering in the upper-right quadrant. In contrast, North American studies showed wider variability and generally lower uptake. Each point corresponds to a specific publication, including: **AUS/NZ** – Mallett (0.39, 0.69), Jayasinghe (0.39, 0.91) **UK/Ireland** – Alkanderi (0.42, 1.00), Elhassan (0.51, 0.97) **North America** – Payne (0.27, 0.66), Bogyo (0.27, 0.91), Pinto e Vairo (0.31, 0.89), Amlie-Wolf (0.33, 0.75), Schott (0.34, 0.93), McFadden (0.37, 0.94), Tan (0.45, 0.97), Lundquist (0.47, 0.83), Thomas 2021 (0.50, 1.00), Bekheirnia (0.51, 0.85), Thomas 2020 (0.60, 0.74), Elliott (0.61, 0.69) **Europe** – Gutierrez (0.26, 0.31), Pode-Shakked (0.57, 0.99), Caliment (0.63, 1.00), Becherucci (0.67, 0.92) All rates are expressed as proportions (0 to 1), based on published percentages. Penetration was defined as the proportion of patients who completed genetic testing out of those eligible and offered testing.

#### Yield vs Test Penetration by Clinic Model

Test penetration was generally high irrespective of clinic model, but yield showed greater dispersion **(Figure 8)**. Multidisciplinary clinics clustered at high penetration (66-100%) with a wide spread of yields—from low (26%) to moderate to high (67%)^5,16,23,25–28,36–38^; a single low-penetration outlier was observed (31%).^26^ Nephrologist-led clinics also demonstrated consistently high penetration (74-100%) with a tighter yield distribution centered in the moderate range (27-60%).^17,18,30–33,35,40^ Evidence for traditional genetics referral and mainstreaming models was limited (single study each) but placed in the mid-to-high penetration with mid-range yield.^19,21^

**Figure 8.**
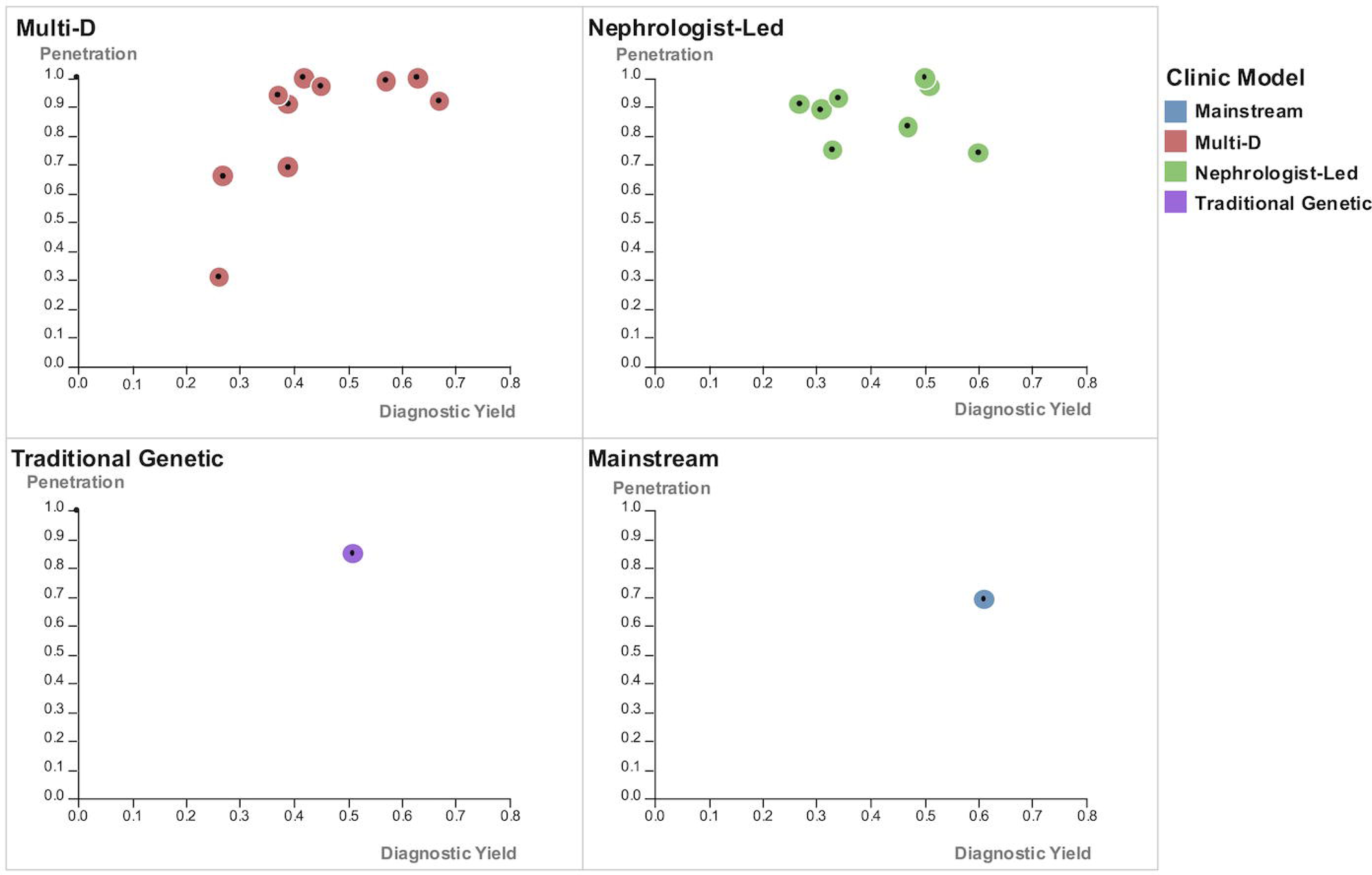
Diagnostic yield versus test penetration in kidney genetics clinics, stratified by clinic model (literature review, n = 20 studies) Scatter plots show diagnostic yield (x-axis) versus genetic test penetration (y-axis), grouped by clinic model. Each dot represents one study and is color-coded by model type: Multidisciplinary (Multi-D, red), Nephrologist-led (green), Traditional genetic referral (purple), and Mainstreaming in general nephrology (blue). **Multi-D clinics** demonstrated consistently high yield and penetration, with some variability: Gutiérrez (0.26, 0.31), Payne (0.27, 0.66), McFadden (0.37, 0.94), Mallett (0.39, 0.69), Jayasinghe (0.39, 0.91), Alkanderi (0.42, 1.00), Tan (0.45, 0.97), Pode-Shakked (0.57, 0.99), Caliment (0.63, 1.00), Becherucci (0.67, 0.92). **Nephrologist-led clinics** showed comparable yield and penetration to Multi-D models: Bogyo (0.27, 0.91), Pinto e Vairo (0.31, 0.89), Amlie-Wolf (0.33, 0.75), Schott (0.34, 0.93), Lundquist (0.47, 0.83), Thomas 2021 (0.50, 1.00), Elhassan (0.51, 0.97), Thomas 2020 (0.60, 0.74). **Traditional genetic referral model** – Bekheirnia (0.51, 0.85). **Mainstream model** – Elliott (0.61, 0.69). All values are expressed as proportions (0 to 1), based on published percentages. Penetration refers to the proportion of eligible patients who completed genetic testing.

#### Yield vs Test Penetration by Testing Approach

Similarly, penetration was uniformly high across testing strategies, while yield varied by approach **(Figure 9)**. Clinics using comprehensive testing (clinical WES/WGS) clustered at high penetration (91-92%) with moderate-to-high yields (39-67%).^23,25^ Phenotypic targeted panels—both stand-alone^18,21,28,30–32,37,38^ and with reflex to WES^5,19,27,33,36^—also showed high penetration (66-100%) with yields in the mid-to-high range (27-63%) and relatively tight clustering. In contrast, hybrid strategies (mix of comprehensive and targeted panels) displayed the greatest dispersion, yield (26-51%), penetration (31-97%)^17,35,40^ including a low-penetration/low-yield outlier.^26^

**Figure 9.**
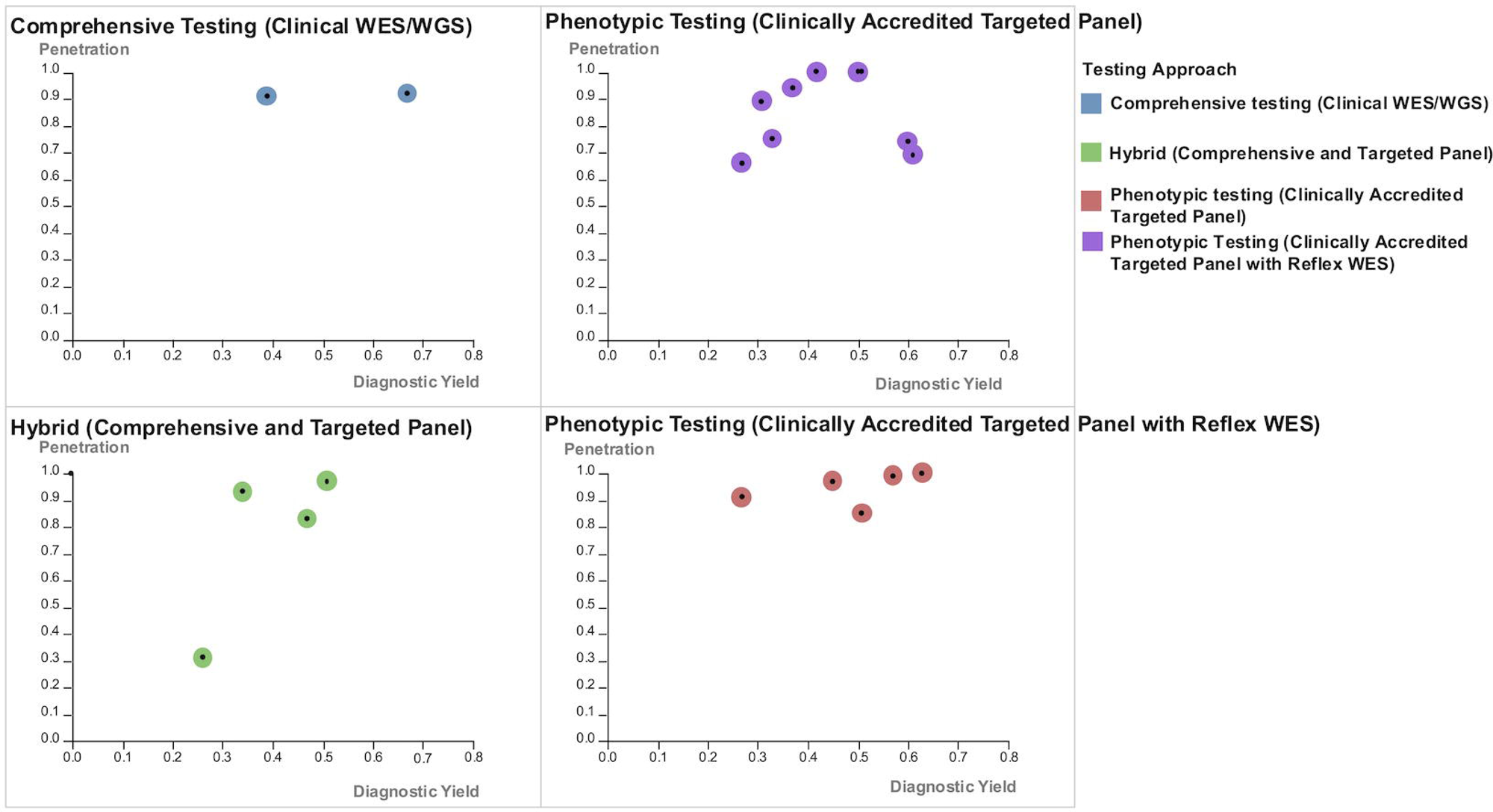
Diagnostic yield versus test penetration in kidney genetics clinics, stratified by testing approach (literature review, n = 19 studies) Scatter plots show diagnostic yield (x-axis) against genetic test penetration (y-axis) for 19 studies that reported both outcomes and testing approaches. Points are grouped by testing strategy and color-coded as follows: blue for comprehensive clinical testing using whole exome or whole genome sequencing (WES/WGS), purple for phenotypic testing using clinically accredited targeted panels, green for hybrid approaches combining comprehensive and targeted panels, and red for phenotypic testing using clinically accredited targeted panels with reflex to WES. No clear difference in yield or penetration was observed between testing strategies, suggesting that patient selection may play a greater role than the specific test used. **Comprehensive WES/WGS** – Jayasinghe (0.39, 0.91), Becherucci (0.67, 0.92) **Phenotypic Targeted Panel Testing** – Payne (0.27, 0.66), Pinto e Vairo (0.31, 0.89), Amlie-Wolf (0.33, 0.75), McFadden (0.37, 0.94), Alkanderi (0.42, 1.00), Thomas 2021 (0.50, 1.00), Thomas 2020 (0.60, 0.74), Elliott (0.61, 0.69) **Hybrid (Comprehensive + Targeted Panel)** – Gutierrez (0.26, 0.31), Schott (0.34, 0.93), Lundquist (0.47, 0.83), Elhassan (0.51, 0.97) **Phenotypic Targeted Panel Testing with Reflex WES** – Bogyo (0.27, 0.91), Tan (0.45, 0.97), Bekheirnia (0.51, 0.85), Pode-Shakked (0.57, 0.99), Caliment (0.63, 1.00) All values are expressed as proportions (0 to 1), based on published percentages. Penetration refers to the proportion of eligible patients who completed genetic testing.

## Discussion

### Geographic Skew of the Evidence Base

This scoping study—including a literature review triangulated with an international stakeholder consultation of clinic leads—maps, for the first time to our knowledge, the global landscape of kidney genetics care across published models, workflows, and outcomes. The evidence base is heavily concentrated in high-income settings (North America, UK/Ireland/EU, and AUS/NZ), with sparse reporting from Asia and little to none from South America and Africa. This pattern likely reflects broader inequities in kidney care capacity and genomics infrastructure in low- and middle-income countries—including workforce shortages,^41,42^ limited access to diagnostics and accredited laboratories, and financial barriers^43^ —and introduces publication and selection biases that limit generalisability. Our international stakeholder consultation findings echo and clarify these gaps, with respondents highlighting constrained resources and reliance on nephrologists to undertake genetics functions in several Asian settings. Critically, populations from under-represented regions account for a large share of the world’s population^44^; their omission from the literature risks perpetuating disparities and narrowing the external validity of current models and outcomes.

### Breadth and Maturity of the Evidence by Domain

We identified seven thematic domains. Most publications cluster in clinic model description/evaluation (n=25), methodology or infrastructure for kidney genomics (n=33), and genomic utility in non-clinic settings (n=26). In contrast, economic evaluations, theory-guided implementation evaluations, stakeholder (patient/clinician) experience, and ethical, legal, and social implications (ELSI) are sparsely represented. These under-studied domains are critical for policy translation into routine care, sustainable service integration, and policy uptake.^45–47^ Among the 25 clinic-model studies, only five were prospective with pre-specified objectives, and qualitative work capturing patient and clinician perspectives was limited. Collectively, this pattern suggests a literature that is rich in descriptions and technical enablers but relatively thin on real-world effectiveness, implementation, equity, and value.

### Clinic Models and Service Architecture

We identified four clinic models in our taxonomy: multidisciplinary, nephrologist-led, mainstreaming within general nephrology, and traditional genetics referral. Multidisciplinary and nephrologist-led models predominated. Multidisciplinary clinics often include nephrologists (adult and/or pediatric), genetic counselors, and clinical geneticists with access to variant review boards. In nephrologist-led services, care is typically anchored by up-skilled nephrologists (genetics-trained or dual-accredited), with embedded genetic counselors and little or no routine clinical geneticist involvement.^48^ Nephrologist-led models emphasise streamlined triage and higher throughput but depend on formal genetics training and decision support. Evidence for mainstreaming remains limited, although several health systems are moving to embed test ordering and counseling within routine nephrology pathways. Recent conceptual frameworks describe multiple mainstreaming configurations that vary by the point at which responsibility is transferred from the non-geneticist clinician to clinical genetics services, underscoring the need to define roles, guardrails, and handovers clearly.^49^

Service maturity appears regionally patterned: early programs were reported in the UK and EU, followed by AUS/NZ, North America, and more recently parts of Asia. Team composition, testing approach, laboratory pathways, and funding mechanisms vary by region, reflecting workforce availability, local capability, and financing models. Testing strategies range from targeted panels to exome/genome sequencing; confirmatory testing and laboratory accreditation pathways (in-house vs commercial, research vs clinically accredited) differ substantially and influence turnaround times, reproducibility, and payer acceptance. Funding models span public reimbursement, institutional support, research fund, insurance, and out-of-pocket payment, with clear implications for penetration and equity.

### Outcomes Beyond Diagnostic Yield

Most reports concentrated on diagnostic utility, with sparse attention to patient- or clinician-reported outcomes and minimal, non-a priori implementation evaluation. This imbalance constrains meaningful conclusions about patient acceptability, clinician adoption, and the sustainability of clinic models, and highlights a critical gap in understanding how these services function beyond diagnostic performance and scalability in real-world health system contexts.

European programs demonstrated the highest diagnostic yield and test penetration, likely attributable to well-established workflows.^27,50^ These typically included standardized referral criteria, deep phenotyping, comprehensive family history collection, rigorous patient and test selection, and integrated multidisciplinary review in both adult and pediatric settings–supported by clearer funding pathways. Such systematic approaches likely enhance both appropriate patient and test selection.

Interestingly, nephrologist-led services performed on par with multidisciplinary clinics when nephrologists were genomics-trained, had embedded genetic counseling, and could access multidisciplinary team input.^17,18,40^ This supports scalable options in settings with limited clinical-genetics capacity, provided training, governance, and decision support are in place.

Yield also appeared relatively independent of test modality. Phenotype-driven targeted gene panels (with or without reflex WES) performed similarly to whole exome or genome sequencing in many studies.^18,19,21,27^ This suggests that meticulous phenotyping and case selection can optimize yield even with more focused testing, although broader sequencing approaches may still offer advantages in detecting structural variants, atypical presentations, and enabling future reanalysis, and gene discovery.

### Implications for Clinical Practice and Service Design

Future efforts should prioritise capacity-building and systematic reporting from under-represented regions to ensure kidney genetics services are adaptable, equitable, and scalable across diverse healthcare systems.^51^ A one-size-fits-all model is unlikely to succeed; instead, programs should strive for contextual fit, explicitly identifying local implementation determinants and aligning clinic models, workforce composition, laboratory infrastructure, and funding pathways with existing health system capabilities and patient case-mix. To advance the field, future evaluations should move beyond retrospective or descriptive studies towards prospective, theory-informed, mixed-methods system-level designs.^52,53^ These should incorporate robust qualitative evaluations of both patient and clinician experiences. Standardised reporting of denominators along the clinical pathway should be routine to enable meaningful benchmarking and cross-model comparisons. Future studies should pre-specify and evaluate key implementation outcomes using established frameworks.^15,54^ Embedding these within hybrid effectiveness-implementation study designs, which assess clinical and implementation outcomes simultaneously, will support a deeper understanding of not just whether kidney genetics clinics diagnose, but how different models perform in delivering scalable, acceptable, and sustainable care in real-world settings.^55^

This will enable future analysis of implementation determinants and the development of tailored strategies for successful kidney genetics clinic models using theory-informed frameworks.^56,57^

### Strengths and Limitations

This study represents the first, to our knowledge, comprehensive synthesis of kidney genetics care, systematically examining clinic models, outcomes, and integrating both empirical evidence from the published literature and experiential data from an international stakeholder consultation of clinic leads. The inclusion of survey data enabled us to capture unpublished or emerging service models and operational practices, enhancing the ecological validity of our findings. By grouping all included studies into seven thematic domains,^13^ developing, and applying a clear taxonomy of clinic models, we were able to compare patterns by region, team composition, and testing approach, and to highlight features associated with higher diagnostic yield and penetration. This triangulation of data sources strengthens the relevance of our recommendations for clinical practice and service design.

Several limitations warrant consideration. The underlying evidence base is heterogeneous, with wide variation in study designs, patient populations, eligibility criteria for genetic testing, types of tests performed, and healthcare system contexts, which limits comparability. Reporting of denominators, time metrics, and outcome definitions was inconsistent, and few studies included patient- or clinician-reported outcomes or theory-informed implementation metrics. Most published studies originated from high-income settings, which may limit generalisability to under-resourced regions. Survey data were self-reported, potentially subject to recall, non-response, and social desirability biases, and regional representation was uneven.

## Conclusion

Current kidney genetics literature and care models are heavily skewed toward high-income countries, with limited representation from under-resourced settings. Traditional genetics referral pathways are declining, while multidisciplinary and nephrologist-led models have gained prominence. The mainstreaming approach is emerging, yet remains under-described. Across all models, clinic structure—including team composition, testing approach, laboratory partnerships, and funding mechanisms—is highly context dependent.

Nephrologist-led models appear comparably effective when supported by adequate training, infrastructure, and governance. Diagnostic yield is more strongly influenced by appropriate patient selection than the specific testing modality used. However, most studies focus narrowly on diagnostic yield, with limited reporting of patient, clinician-reported, or implementation outcomes.

Moving forward, kidney genetics services must be tailored to local system-level determinants, incorporating context-specific strategies to address barriers and enablers. Standardised reporting of diagnostic pathways, testing penetration, and time-to-diagnosis is critical for benchmarking. Theory-informed, prospective implementation evaluations will be essential to understand not just whether models work, but how they deliver equitable, acceptable, and sustainable care. These implementation determinants and matched strategies should be explored in greater detail to inform future pathways to optimal benefit for patients and families globally.

## Data Availability

This study includes data from publicly available sources and an international expert consultation of kidney genetics clinic leads. All data extracted from published studies are available within the article and supplementary materials. The survey data are not publicly available due to stakeholder confidentiality but may be shared in de-identified form upon reasonable request to the corresponding author, contingent upon institutional approval and a data use agreement.

## Acknowledgements

The authors thank all international kidney genetics clinic leads who generously contributed their time and insights to the consultation. A complete list of the Global Kidney Genetics Clinic Leads Collaborators (n=48), including names, affiliations, and countries, is provided in the **Supplemental Table 10**.

We also acknowledge Louisse RG Lim and An Ni Lim for their valuable contributions to data visualization for this study.

This work is part of the GENEKIDS-PRO research program.

RSL is supported by the Singapore Ministry of Health’s National Medical Research Council (NMRC) Clinician Scientist Individual Research Grant New Investigator Grant (CNIG24jul-0012) and the NMRC Research Training Fellowship (RTF24jul-0007).

AJM is supported by a Queensland Health Advancing Clinical Research Fellowship.

## Author Contributions

RSL, NS, AJM, ECCY, CP, EB, and SA conceived and designed the study. RSL performed the data analysis and data visualisation and drafted the manuscript. CP, NS, and AJM critically revised the manuscript for important intellectual content. All authors reviewed and approved the final manuscript.

**Supplementary material is available online at** www.kidney-international.org.

---

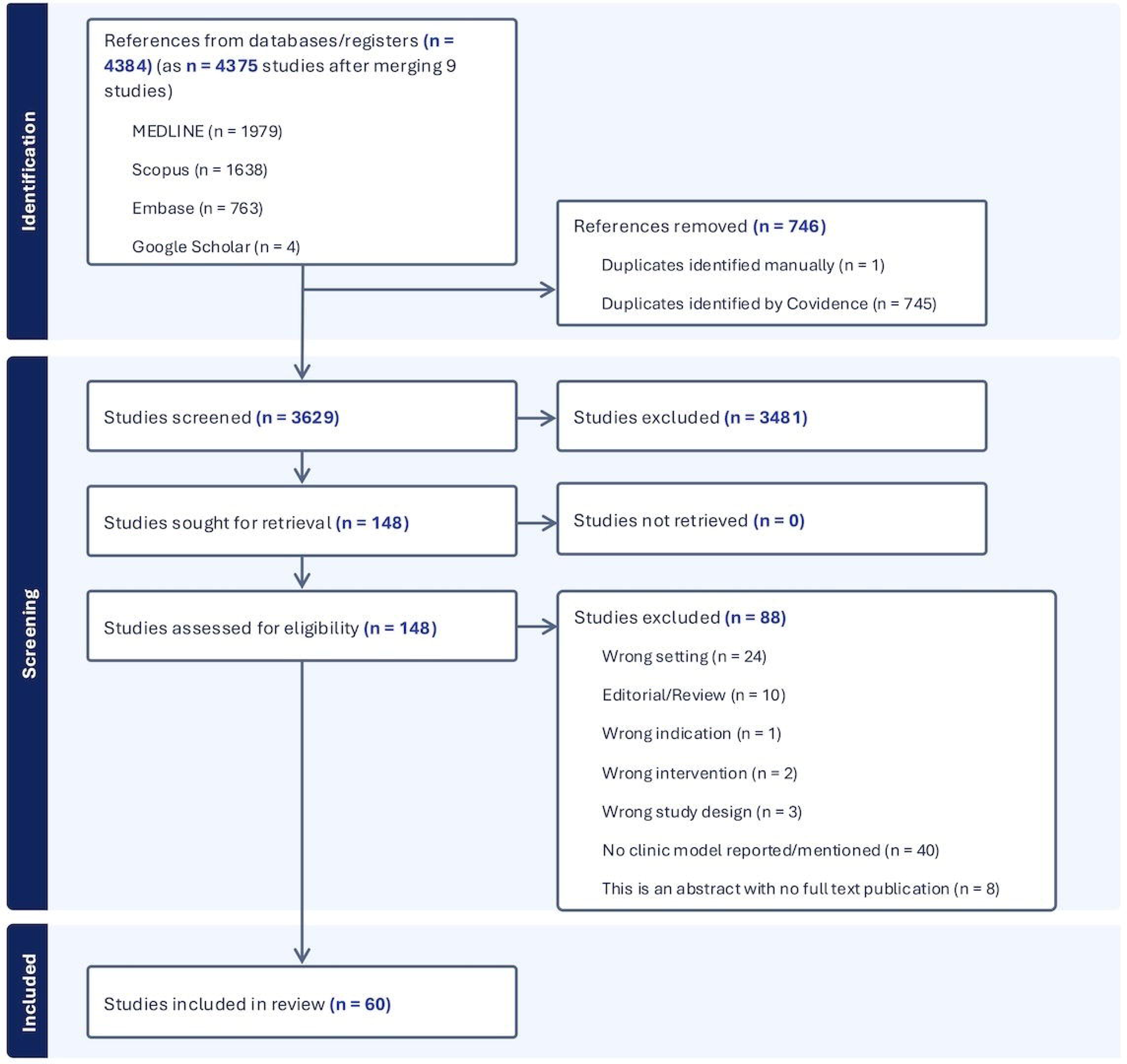
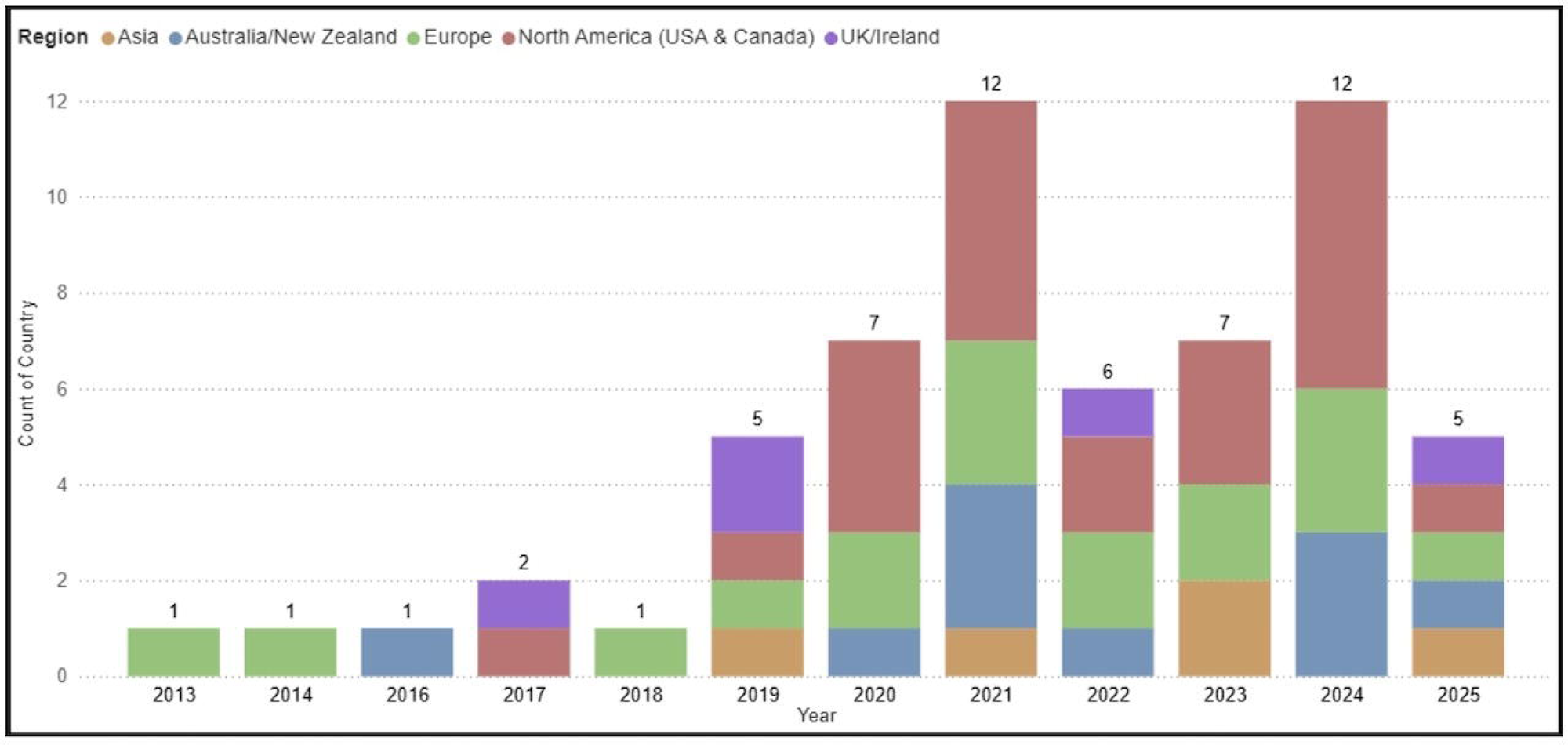
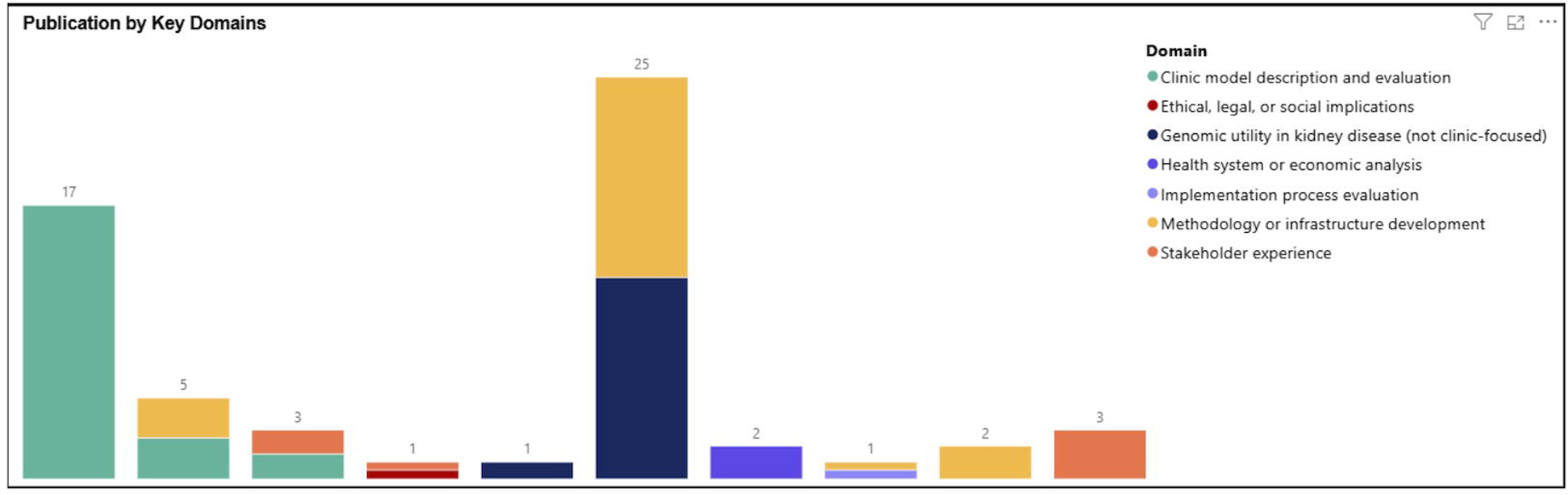
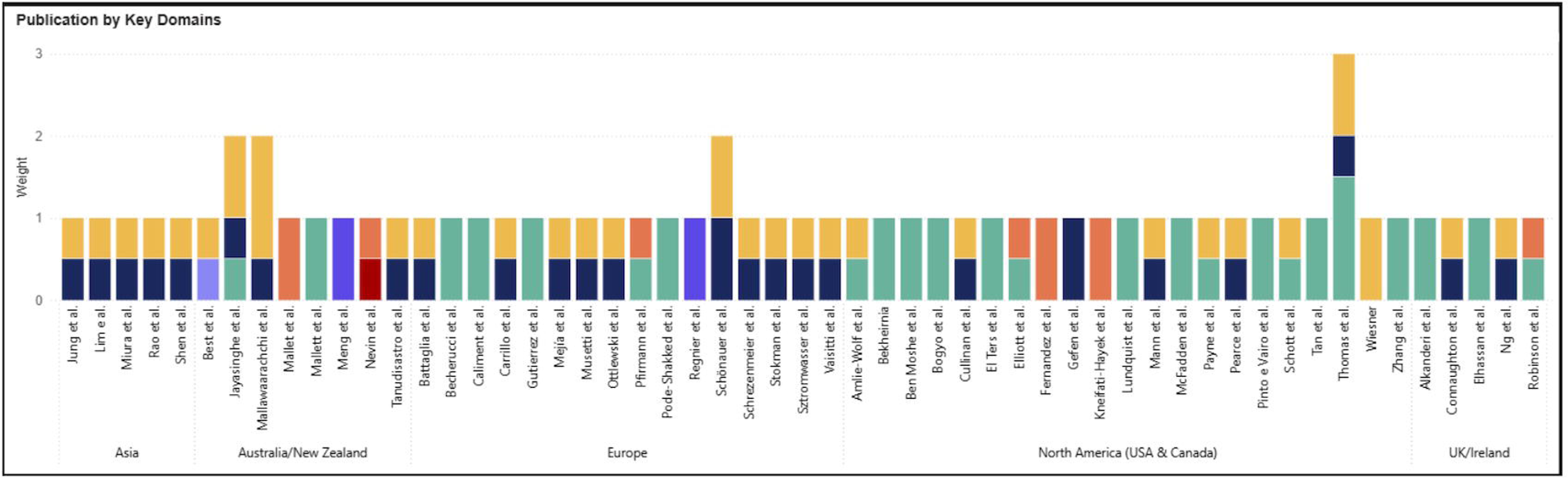

