## Supplementary Material for "Defining the Global Landscape of Kidney Genetics Care— A Scoping Review and International Stakeholder Consultation of Clinic Models and Outcomes"

| **Contents** | **Page** |
| --- | --- |
| Supplemental Table 1: Preferred Reporting Items for Systematic reviews and Meta-Analyses extension for Scoping Reviews (PRISMA-ScR) Checklist | 2-3 |
| Supplemental Table 2: Search strategies and keywords used across MEDLINE (PubMed), Embase, and Scopus for identifying studies on kidney genetics service | 4 |
| Supplemental Table 3: List of data extraction fields | 5 |
| Supplemental Table 4: Taxonomy of kidney genetics clinic models | 6-7 |
| Supplemental Table 5: Survey questionnaires | 8-11 |
| Supplemental Table 6: Domain 2 and 3 (Genomic utility in kidney disease in non-clinic settings, methodology, or infrastructure development to support the implementation of kidney genomics, n=28) | 12-16 |
| Supplemental Table 7: Domain 4 (Stakeholder experience, n=4) | 17-18 |
| Supplemental Table 8: Domain 5 (Health economic and health system analysis, n=2) | 19 |
| Supplemental Table 9: Domain 6 (Kidney genomic implementation process evaluation, n=1) | 20 |
| Supplemental Table 10: List of Global Kidney Genetics Clinic Leads Collaborators (n = 48) | 21-22 |
| Supplemental Figure Legends | 23-25 |
| Supplemental References | 26-28 |

**Supplemental Table 1: Preferred Reporting Items for Systematic reviews and Meta-Analyses extension for Scoping Reviews (PRISMA-ScR) Checklist**

| **SECTION** | **ITEM** | **PRISMA-ScR CHECKLIST ITEM** | **REPORTED ON PAGE #** |
| --- | --- | --- | --- |
| **TITLE** | | | |
| Title | 1 | Identify the report as a scoping review. | 1 |
| **ABSTRACT** | | | |
| Structured summary | 2 | Provide a structured summary that includes (as applicable): background, objectives, eligibility criteria, sources of evidence, charting methods, results, and conclusions that relate to the review questions and objectives. | 3-4 |
| **INTRODUCTION** | | | |
| Rationale | 3 | Describe the rationale for the review in the context of what is already known. Explain why the review questions/objectives lend themselves to a scoping review approach. | 5 |
| Objectives | 4 | Provide an explicit statement of the questions and objectives being addressed with reference to their key elements (e.g., population or participants, concepts, and context) or other relevant key elements used to conceptualize the review questions and/or objectives. | 5 |
| **METHODS** | | | |
| Protocol and registration | 5 | Indicate whether a review protocol exists; state if and where it can be accessed (e.g., a Web address); and if available, provide registration information, including the registration number. | 4 |
| Eligibility criteria | 6 | Specify characteristics of the sources of evidence used as eligibility criteria (e.g., years considered, language, and publication status), and provide a rationale. | 5 |
| Information sources* | 7 | Describe all information sources in the search (e.g., databases with dates of coverage and contact with authors to identify additional sources), as well as the date the most recent search was executed. | 6 |
| Search | 8 | Present the full electronic search strategy for at least 1 database, including any limits used, such that it could be repeated. | S4 |
| Selection of sources of evidence† | 9 | State the process for selecting sources of evidence (i.e., screening and eligibility) included in the scoping review. | 6 |
| Data charting process‡ | 10 | Describe the methods of charting data from the included sources of evidence (e.g., calibrated forms or forms that have been tested by the team before their use, and whether data charting was done independently or in duplicate) and any processes for obtaining and confirming data from investigators. | 6-7 |
| Data items | 11 | List and define all variables for which data were sought and any assumptions and simplifications made. | S5 |
| Critical appraisal of individual sources of evidence§ | 12 | If done, provide a rationale for conducting a critical appraisal of included sources of evidence; describe the methods used and how this information was used in any data synthesis (if appropriate). | NA |
| Synthesis of results | 13 | Describe the methods of handling and summarizing the data that were charted. | 7 |
| **RESULTS** | | | |
| Selection of sources of evidence | 14 | Give numbers of sources of evidence screened, assessed for eligibility, and included in the review, with reasons for exclusions at each stage, ideally using a flow diagram. | 9 |
| Characteristics of sources of evidence | 15 | For each source of evidence, present characteristics for which data were charted and provide the citations. | 23-25 |
| Critical appraisal within sources of evidence | 16 | If done, present data on critical appraisal of included sources of evidence (see item 12). | NA |
| Results of individual sources of evidence | 17 | For each included source of evidence, present the relevant data that were charted that relate to the review questions and objectives. | 9-13 |
| Synthesis of results | 18 | Summarize and/or present the charting results as they relate to the review questions and objectives. | 9-13 |
| **DISCUSSION** | | | |
| Summary of evidence | 19 | Summarize the main results (including an overview of concepts, themes, and types of evidence available), link to the review questions and objectives, and consider the relevance to key groups. | 14-16 |
| Limitations | 20 | Discuss the limitations of the scoping review process. | 6-17 |
| Conclusions | 21 | Provide a general interpretation of the results with respect to the review questions and objectives, as well as potential implications and/or next steps. | 17 |
| **FUNDING** | | | |
| Funding | 22 | Describe sources of funding for the included sources of evidence, as well as sources of funding for the scoping review. Describe the role of the funders of the scoping review. | 18 |

JBI = Joanna Briggs Institute; PRISMA-ScR = Preferred Reporting Items for Systematic reviews and Meta-Analyses extension for Scoping Reviews.

* Where *sources of evidence* (see second footnote) are compiled from, such as bibliographic databases, social media platforms, and Web sites.

† A more inclusive/heterogeneous term used to account for the different types of evidence or data sources (e.g., quantitative and/or qualitative research, expert opinion, and policy documents) that may be eligible in a scoping review as opposed to only studies. This is not to be confused with *information sources* (see first footnote).

‡ The frameworks by Arksey and O’Malley (6) and Levac and colleagues (7) and the JBI guidance (4, 5) refer to the process of data extraction in a scoping review as data charting*.*

§ The process of systematically examining research evidence to assess its validity, results, and relevance before using it to inform a decision. This term is used for items 12 and 19 instead of "risk of bias" (which is more applicable to systematic reviews of interventions) to include and acknowledge the various sources of evidence that may be used in a scoping review (e.g., quantitative and/or qualitative research, expert opinion, and policy document).

**Supplemental Table 2: Search strategies and keywords used across MEDLINE (PubMed), Embase, and Scopus for identifying studies on kidney genetics service**

| **Database** | **Subject headings** | **Keywords** |
| --- | --- | --- |
| MEDLINE (PubMed)  Using MeSH terms | Kidney Diseases OR Nephrology | Kidney Dise* OR Nephrol* |
|  | Genetic Testing OR Genetics OR Genetic Services OR Genetic Counseling | Genet* OR Genomic* |
|  | Health Services OR Patient Care Team | Health* Serv* OR Services, Health OR Health Care Team OR Healthcare Team OR Interdisciplinary Health Team OR Medical Care Team OR Multidisciplinary Care Team OR Multidisciplinary Health Team OR Kidney Genet* Clinic* OR Renal Genet* Clinic* |
| Embase  Using Emtree terms | Kidney Disease OR Nephrology | Kidney Disease* OR Kidney Disorder* OR Renal Disease* OR Nephrol* OR Genetic Kidney Disease* OR Monogenic Kidney Disease* OR Inherited Kidney Disease* OR Hereditary Kidney Disease* |
|  | Genetic Service OR Genetic Screening OR Genetic Counseling OR Genetic Disorder OR Genomics | Genetic Service* OR Genetic Screen* OR Genetic Test* OR Screen*, Genetic OR Genetic Test* OR Genetic Counsel* OR Counsel*, Genetic OR Genetic* OR Genetic Disease* OR Genomic* |
|  | Multidisciplinary Team OR Collaborative Care Team OR Health Service OR Patient Care Team | Inter-disciplinary Team OR Interdisciplinary Team OR Multi-disciplinary Team OR Inter-disciplin* OR Interdisciplin* OR Multi-disciplin* OR Multidisciplin* OR collaborative health care team OR collaborative healthcare team OR collaborative patient care team OR Collaborat* OR Health Care Service OR Health Service* OR Healthcare Service* OR Patient Health Care Team OR Patient Healthcare Team |
| Scopus |  | ( TITLE-ABS  KEY ( kidney OR renal AND disease* ) AND TITLE-ABS-KEY ( genetic AND * OR genomic* ) AND TITLE-ABS-KEY ( multidisciplinary OR interdisciplinary OR service OR care OR clinic ) ) |

**Supplemental Table 3: List of data extraction fields**

| **Data Category** | **Data Fields under this Category** |
| --- | --- |
| 1. Study characteristics | Country of publication  First author  Year of publication  Study design  Study period  Study aims  Study methodology  Clinical setting and healthcare context  Study population and sample size  Inclusion and exclusion criteria |
| 2. Clinic model description | Clinic model type  Clinic team members  Clinic workflow description |
| 3. Genomic testing approach | Test type  Number of patients sequenced  Genetic lab type  Funding mechanism |
| 4. Outcomes reported | Service outcomes   i. Diagnostic yield ii. Turn-around-time   Patient reported outcome measures  Implementation outcomes |

**Supplemental Table 4: Taxonomy of kidney genetics clinic models**

| **Domains** | **Subdomains** | **Definitions** |
| --- | --- | --- |
| 1: Clinic Model  (Service Design) | 1a: Multidisciplinary Integrated Kidney Genetics Clinic | Joint clinic co-managed by nephrologists and the genetics team, with shared responsibility for genetic test ordering and patient care. Typically delivered in a concurrent or tightly coordinated format. |
|  | 1b: Genetic-Trained Nephrologist-Led Clinic with Genetics Team Support | Nephrologist with genetics training leads care and test ordering, supported by genetic counselor and/or clinical geneticist via virtual, asynchronous, or consultative input. |
|  | 1c: Mainstreaming in General Nephrology Clinic | General nephrologist independently initiates genetic testing and provides basic counseling. No formal involvement from the genetics team. |
|  | 1d: Traditional Genetic Referral Model | Nephrologist refers patient to a separate clinical genetics service. Test ordering and counseling are handled independently by the genetics team. |
| 2: Team Composition and Leadership | 2a: Genetic-Trained Nephrologist-Led | Clinic is led by a nephrologist with genetics training, who oversees genomic testing and counseling. May be supported by a genetic counselor and/or clinical geneticist. |
|  | 2b: Geneticist-Led | Clinic is led by a clinical geneticist or genetic counselor. Nephrologist contributes renal phenotyping and clinical input as needed. |
|  | 2c: Nephrologist-Geneticist Jointly Led (Multidisciplinary Team) | Nephrologist, genetic counselor, and clinical geneticist share leadership and clinical decision-making responsibilities. |
|  | 2d: General Nephrologist-Led | General nephrologist leads the clinic and coordinates genetic testing and counseling without formal genetics team involvement. |
| 3: Test Ordering Authority | 3a: Genetics-Trained Nephrologist Orders Test | Genetics-trained nephrologist independently initiates and manages genetic testing. |
|  | 3b: Geneticist Orders Test | Test is initiated solely by the clinical genetics team (clinical geneticist and/or genetic counselor). |
|  | 3c: Shared Test Ordering | Test decision and ordering are made collaboratively between nephrology and genetics teams. |
|  | 3d: General Nephrologist Orders Test | General nephrologist independently orders genetic testing without formal genetics input. |
| 4: Clinic Setting/Session Format | 4a: Concurrent Multidisciplinary Team | Patient seen jointly by nephrologist and genetics providers in the same session. |
|  | 4b: Sequential (same day) | Patient seen by nephrology and genetics providers separately but on the same day. |
|  | 4c: Sequential (different days) | Nephrology and genetics visits occur on different days, scheduled independently. |
|  | 4d: Nephrology-Only Clinic | All care delivered within nephrology; no formal genetics provider involvement. |
|  | 4e: Genetic Clinic | Patient seen in a genetics service, independent of nephrology clinic. |
| 5: Referral Criteria (Patient Inclusion Criteria) | 5a: Phenotype-Based (broad categories) | Referral or inclusion is based on clinical presentation across a broad spectrum of suspected genetic kidney disease phenotypes (e.g., cystic, glomerular, tubulointerstitial, nephrolithiasis). |
|  | 5b: Phenotype-Based (narrow/single category) | Referral or inclusion is based on a narrow or single suspected phenotype (e.g., glomerular or cystic only). |
|  | 5c: Universal Screening | All patients with chronic kidney disease are offered genetic testing, regardless of phenotype or clinical presentation. |
|  | 5d: Protocol-/Algorithm-Based | Referral is triggered by predefined protocols or clinical decision support algorithms. |
| 6. Genetic Testing Approach | 6a: Phenotype-Driven Testing Using Targeted Panel ± Reflex WES/WGS | Clinically accredited phenotype-based targeted panel is used initially, with optional reflex to WES or WGS if panel testing is inconclusive. |
|  | 6b: Comprehensive Clinical WES/WGS ± analysis of virtual panel(s) | Clinical whole exome or genome sequencing is used as first-line, often with virtual panels applied for phenotype-guided analysis. |
|  | 6c: Primary Research Testing with Clinical Confirmation | Initial testing is performed in a research setting, with clinically accredited confirmatory testing if a candidate variant is identified. |
|  | 6d: Hybrid Approach | Combination of clinical and research testing strategies or multi-step workflows adapted to available resources and case complexity. |
| 7. Genetic Counselling | 7a: Counselling by Genetic Counselors | Genetic counsellor provides both pre- and post-test counselling. |
|  | 7b: Counselling by Clinical Geneticist | Clinical geneticist provides both pre/post-test counselling |
|  | 7c: Counselling by Nephrologist | Nephrologist independently provides all counselling. |
|  | 7d: Shared Counselling | Counselling is shared between nephrologist and genetic counsellor and/or clinical geneticist, depending on clinic context. |
| 8. Genetic Laboratory | 8a: In-House Accredited Genetic Laboratory | Genetic testing performed in a clinically accredited laboratory within the same institution or health system. |
|  | 8b: Commercial Accredited Genetic Laboratory | Testing outsourced to a commercially run, clinically accredited genetic laboratory. |
|  | 8c: Research Non- Accredited Genetic Laboratory | Testing performed in a research laboratory that is not clinically accredited; results may require confirmatory testing. |
| 9. Cascade Testing Model | 9a: Active Outreach and Facilitated Family Testing | Clinic actively engages at-risk relatives, facilitates coordination and logistics for cascade testing. |
|  | 9B: Patient-Initiated Family Testing with Guidance | Index patient informs relatives and testing is arranged based on guidance or resources provided by the clinic. |
|  | 9C: Referred Family Testing Via Genetic Services | Index patient or family is referred to external clinical genetics services for cascade testing. |
| 10: Funding Model | 10a: Clinical | Services are reimbursed through routine clinical billing, public health system, or national health insurance. |
|  | 10B: Research | Services are supported by research grants or pilot programs, often requiring patient participation in studies. |
|  | 10C: Hybrid | A combination of clinical and research funding streams supports service delivery. |
|  | 10D: Philanthropy/Non-governmental organizations | Services funded by philanthropic contributions, charity foundations, or non-governmental organizations. |

*This taxonomy was iteratively developed through literature review and expert consultation, and subsequently applied to both included studies and survey responses. Definitions reflect real-world variations in service design, team composition, and testing approaches.*

**Supplemental Table 5: Survey questionnaires**

| **Domains** | **Questions** |
| --- | --- |
| 1. Demography | a. Country of practice  b. Institution/organization  c. What is your current primary role in your kidney genetics clinic or service?   i. Nephrologist (Adult) ii. Nephrologist (Pediatric) iii. Clinical Geneticist iv. Genetic Counselor v. Laboratory Geneticist vi. Researcher/Academic vii. Service Manager   d. How many years have you been involved in kidney genetics clinical care or service development?   i. Less than 1 year ii. 1-3 year iii. 4-6 years iv. 7-9 years v. 10 years or more   e. What best describes your level of involvement in your kidney genetics service?   i. I lead or co-lead the clinic/service ii. I am part of the core multidisciplinary team I support the service (e.g. admin, lab, coordination) iii. Other:   f. How frequently are you involved in kidney genetics clinic activities?   i. Weekly ii. Fortnightly iii. Monthly iv. Occasionally (As needed) v. Rarely / Not currently involved |
| 2. Overview of Your Kidney Genetics Clinic/Service | a. What is the official name of your kidney genetics clinic/service?  b. In what year was your kidney genetics clinic/service established?  c. Where is your service based?   i. Tertiary hospital ii. Community hospital iii. Academic or university medical centre iv. Private practice or specialist clinic v. Regional genetics centre vi. Other:   d. How is your service delivered?   i. In-person clinics ii. Virtual/telehealth clinics iii. Hybrid (in-person + virtual) iv. Inpatient consults v. Outreach (e.g. regional sites or satellite clinics)   e. What is the approximate frequency of your kidney genetics clinics?   i. Weekly ii. Fortnightly iii. Monthly iv. Ad hoc / On demand v. Other:   f. How would you describe your clinic model?   i. Multidisciplinary Integrated clinic ii. Genetic-trained nephrologist-led genetics clinic assisted by genetic team iii. General nephrology clinic (mainstream) iv. Genetic-referral based model v. Other:   g. What best describe the clinical genetic testing and counselling pathway in your clinic model?   i. Integrated genetic testing and counselling within multidisciplinary kidney genetics clinic ii. Genetic trained nephrologist-led genetic test ordering and counselling assisted by genetic team iii. General nephrologist-led genetic test ordering and counselling under routine outpatient setting iv. Genetic test ordering and counselling by external genetic service   h. What best describe your kidney genetics clinic service setting?   i. Multidisciplinary team in one setting running concurrently ii. Multidisciplinary team in one setting running sequentially iii. Multidisciplinary team in different setting iv. Nephrology clinic with genetic testing v. Traditional nephrology clinic with referral to external genetics service vi. Other:   i. How would you describe your kidney genetics clinic service delivery?   i. Clinical service based ii. Research based iii. Hybrid (clinical and research based)   j. Team composition: which disciplines are routinely involved in your kidney genetics clinic? (select all that apply)   i. Nephrologist (Adult) ii. Nephrologist (Pediatric) iii. Clinical geneticist iv. Genetic counsellors v. Renal genetic nurse vi. Laboratory geneticist/scientist vii. Researcher viii. Social worker/psychologist ix. Other: |
| 3. Patient Population and Referral Pathway | a. What types of patients are seen in your clinic?   i. Adults ii. Children iii. Both |
| 4. Genetic Counseling and Testing | a. Who provides pre and post-test genetic counselling in your clinic? (select all that apply)   i. Genetic counsellor ii. Clinical geneticist iii. Nephrologist iv. Nurse v. Hybrid depending on case complexity vi. Limited/No genetic counselling   b. What is your approach to genetic testing?   i. Phenotypic testing using clinically accredited targeted panel with reflex WES/WGS testing ii. Comprehensive testing using WES/WGS [with analysis of virtual panel(s)] iii. Primary research testing with reflex clinical testing confirmation iv. Hybrid v. Other:   c. Where is genetic testing performed? (select all that apply)   i. In-house accredited genetic laboratory ii. Commercial accredited genetic laboratory iii. Research non-accredited genetic laboratory iv. Other: |
| 5. Cascade Testing and Family Management | a. Do you offer cascade testing to family members of probands?   i. Yes, routinely ii. Yes, on request iii. No iv. Not applicable   b. Who coordinates or facilitates family cascade testing? (select all that apply)   i. Genetic counselors ii. Clinical geneticist iii. Nephrologist iv. Nurse or coordinator v. Family initiates testing independently vi. Other: |
| 6. Service Funding and Sustainability | a. How is your kidney genetics clinic funded? (select all that apply)   i. Public health system ii. Institutional funding iii. Philanthropic or research grant iv. Private billing / out-of-pocket Insurance v. Other:   b. How is genetic testing funded for patients? (select all that apply)   i. Publicly subsidised ii. Insurance-covered iii. Out-of-pocket by patient iv. Research grant v. Mixed model vi. Other: |

**Supplemental Table 6: Domain 2 and 3 (Genomic utility in kidney disease in non-clinic settings, methodology, or infrastructure development to support the implementation of kidney genomics, n=28)**

| **Study (Author, Year)** | **Country** | **Study Design and Setting** | **Clinical Pathway/**  **Methodology/Infrastructure Studied** | **Team Composition** | **Referral Criteria** | **Patient Population** | **Genetic Testing Approach** | **Diagnostic Yield (%)** | **Test Uptake Rate (%)** |
| --- | --- | --- | --- | --- | --- | --- | --- | --- | --- |
| Tanudisastro et al., 2021 ^1^ | AUS/NZ | Retrospective  cohort  Single tertiary center (Sydney) | Centralized Accredited Australia and New Zealand Renal Gene Panels **clinical laboratory service** | Nephrologist  Clinical geneticist  Laboratory scientist | Phenotype based broad spectrum | Adults  (n=271) and children (n=281) | Comprehensive testing using clinical WES with analysis of virtual panels | 189/542 family unit (35%) | 552/552 patients (100%) |
| Mallawaarachchi et al., 2024 ^2^ | AUS | Prospective cohort  National multi-center | Accredited **whole genome sequencing with tier-based broad analysis** of kidney, human, and mitochondrial disease genes | Distributed kidney genetics clinic network model supported by a centralized MDT | Unexplained stage 5 CKD at the age of 50 years or less. | Adult  and children (n=100) | Comprehensive testing using clinical WGS with analysis of virtual panels | 25/100 (25%) | 100/145 (69%) |
| Jayasinghe et al., 2024 ^3^ | AUS | Retrospective and prospective cohorts  National multi-center | National Multidisciplinary Kidney Genetics **Clinic Network** | Distributed kidney genetics clinic network model supported by a centralized MDT | Phenotype based broad spectrum | Adults  (n=849) and children (n=532) | Hybrid | 606/1322 (45.8%) | 1381/1506 (92%) |
| Mallawaarachchi et al., 2025 ^4^ | AUS | Retrospective and prospective cohort  National multi-center | Comprehensive advanced research analytics with **functional genomics approaches** | National multidisciplinary genomic research collaborative | Patients with suspected unexplained genetic kidney disease who remained unsolved after standard genomic testing. | Ongoing | Hybrid  Short read WGS  RNA sequencing  Long read sequencing  Functional genomics | NA | NA |
| Ng et al., 2025 ^5^ | UK | Retrospective cohort  Single tertiary center | Genetic **testing protocol** using 8-gene next generation sequencing panel | Pediatric nephrologist | Children with:  1. Persistent microscopic hematuria was defined as  ≥10× 106/L red  blood cells persisting for>6 months. OR  2. Testing in a proband with hematuria when  (i) A first degree relative with hematuria of unexplained CKD; or  (ii) Electron microscopy evidence of Alport syndrome; or  (iii) Clinical features of Alport syndrome (sensorineural hearing loss, perimacular flecks or anterior lenticonus). | Children (n=224)  134 had genetic testing | Accredited targeted panel | 91/134 (68%) | NA |
| Connaughton et al., 2019 ^6^ | Ireland | Prospective cohort  Multi-center | Genetic testing using **tiered phenotype-driven gene prioritization approach** | NR | Adult CKD patients with:  i) Positive family history of CKD OR  (ii) Extra-renal features | Adults (n=138)  114 families | Comprehensive testing using clinical WES with analysis of virtual panels | 42/114 families (37%) | NR |
| Thomas et al., 2017 ^7^ | USA | Prospective pilot  Single tertiary center | **Genetic testing** using comprehensive 115-gene panel | NR | Living related kidney donors in transplant candidates with suspected genetic kidney disease | 4 transplant candidates and 6 related living kidney donors | Accredited targeted panel | Transplant candidate ¾ (75%)  Donor ¾ (75%) screened negative and accepted for donation | NR |
| Mann et al., 2019 ^8^ | USA | Retrospective cohort  Single tertiary center | **Genetic testing** using research WES | NR | Kidney transplant recipient with CKD onset before 25 years of age | Children (n=104) | Research WES | 34/104 (32.7%) | NR |
| Wiesner et al., 2020 ^9^ | USA | Cross-sectional  Muti-center | **Approaches for the return of genomic results** to participants and their healthcare providers to identify “best practices” within the healthcare system. | NR | Patients enrolled to Electronic Medical Records and Genomics (eMERGE3) Network study | Adult and children | Comprehensive testing using clinical WES with analysis of virtual panels | NA | NA |
| Gefen et al., 2023 ^10^ | USA | Retrospective cross-sectional  Multi-center | **Genetic testing** using 40-gene nephrolithiasis panel testing | NR | Children age 21 years or less with presence of nephrolithiasis and/or nephrocalcinosis on diagnostic imaging or evidence of a passed stone | Children (n=113) | Accredited targeted panel | 13/113 (11.5%) | NR |
| Pearce et al., 2024 ^11^ | USA | Retrospective cohort  Single tertiary center | **Genetic** **testing** using nephrolithiasis 35-gene panel testing | NR | Adult patients with nephrolithiasis age onset </=18, OR elevated urinary oxalate levels >40mg/day. | Adults (n=36) | Accredited targeted panel | 5/36 (14%) | NR |
| Cullinan et al., 2020 ^12^ | Canada | Retrospective cohort  Single center | **Genetic** **e-referral decisional algorithm** specific to Wilm’s Tumour to identify children at elevated risk of having a cancer predisposition syndrome | Geneticist | Children aged <18 diagnosed with and/or treated for Wilm’s Tumour | Children (n=107) | Accredited targeted panel | 16/101 (15.8%) | 101/107 (94%) |
| Musetti et al., 2014 ^13^ | Italy | Retrospective cohort  Single center | ***HNF1B*** **genetic** **screening criteria** | NR | Adult patients with unexplained CKD with renal structure abnormalities or positive family history of nephropathy | Adult (n=70) | Sanger sequencing combined with MLPA | 6/67 (9%) | 67/70 (96%) |
| Vaisitti et al., 2021 ^14^ | Italy | Retrospective cohort  Multi-center | **Regional web-based multidisciplinary genetic service** to provide pre-test genetic counselling and consultation to determine eligibility of genetic testing | Nephrologist  Geneticist | Patients with unexplained CKD | Adults (n=86) and children (n=52) | Comprehensive testing using clinical WES with analysis of virtual panels | 78/138 (56.5%) | 138/138 (100%) |
| Battaglia et al., 2025 ^15^ | Italy | Prospective cohort  Multi-center | **Expanded Fabry’s disease genetic screening approach** in out-patient nephrology clinics | NR | Adult non-dialysis CKD stages 1-5 patients.  Genetic testing for fabry’s disease was performed in all females regardless of enzyme activity and in males with low α-Gal A activity. | Adults (n=173) | Sanger sequencing | 10/183 (5.4%)  Proband: 4/173 (2.2%)  Family members: 6/10 (60%) | 173/173 (100%) |
| Mejía et al., 2013 ^16^ | Spain | Retrospective and prospective cohort  Multi-center | **Tubulopathy network-**based registry and database | Nephrologist  Geneticist  Molecular biologist | Patients with suspected genetic tubulopathy | Adult and children  Total (n=222) | Clinically accredited targeted panel with MLPA in inconclusive cases | 26/44 (59%) | NR |
| Galán Carrillo et al., 2024 ^17^ | Spain | Retrospective cohort  Multi-center | **Multidisciplinary Unit** for Hereditary Kidney Diseases of the Region of Murcia (MUHKD-RM) | Nephrologist  Clinical geneticist  Molecular geneticist | Patients who had a genetic study from the beginning of the NGS panel of hereditary kidney disease implementation | Adult and children  Total (n=360) | Clinically accredited targeted panel with reflex clinical exome or MLPA in inconclusive cases | 120/360 (33.3%) | NR |
| Ottlewski et al., 2019 ^18^ | Germany | Cross sectional  Single center | **Genetic testing** using comprehensive 209-gene panel covering diverse renal phenotypes | NA | Kidney transplant waitlist patients with kidney failure of undetermined etiology. | Adult (n=57) | Clinically accredited targeted panel with MLPA to confirm CNV | 6/50 (12%) | 50/57 (88%) |
| Schönauer et al., 2020 ^19^ | Germany | Prospective cohort  Single center | **Genetic testing** using tier-base extended genetic diagnostics modalities | NA | Patients with clinically diagnosed ADPKD | 100 families | Clinically accredited targeted panel, followed by MLPA for CNV, and exome sequencing in consecutively unsolved cases. | 82/100 families (82%) using panel testing with MLPA. 2/9 (22%) using exome. | NR |
| Schrezenmeier et al., 2021 ^20^ | Germany | Cross-sectional  Single center | **Genetic testing** using comprehensive 600-gene panel covering diverse renal phenotypes | NA | Kidney transplant waitlist patients with kidney failure of undetermined etiology:  1. Onset of kidney failure <40 years old  2. FSGS or aHUS proven on kidney biopsy of any age. | Adult (n=137) | Clinically accredited targeted panel | 14/126 (11.1%) | 126/137 (92%) |
| Schönauer et al., 2022 ^21^ | Germany | Prospective cohort  Single center | **Genetic testing** using targeted kidney stone 45-gene panel | NA | Any adults admitted for kidney stone disease intervention | Adult (n=276) | Clinically accredited targeted panel | 16/236 (6.8%) | 236/276 (86%) |
| Stokman et al., 2018 ^22^ | The Netherlands | Retrospective cohort  Multi-center | National **nephronophthisis-related ciliopathies (NPH-RC) registry** | Nephrologist  Clinical geneticist | Patients with molecularly confirmed diagnosis of NPH-RC or suspected NPH-RC as defined as:  1. CKD stage 2-5  2. Family history compatible with autosomal recessive inheritance and  3. Extrarenal features associated with a ciliopathy and/or clinical characteristics of NPH | n=40  Known molecular diagnosis (n=23)  Unknown molecular diagnosis (n=17) | Hybrid  Clinically accredited targeted panel (15 genes)  MLPA  WES | 5/13 (38%) | 13/17 (76%) |
| Sztromwasser et al., 2020 ^23^ | Poland | Retrospective cross-sectional  Multi-center | **Refined clinical scoring system** to improve the diagnostic yield of *HNF1B* genetic testing | NA | Patients tested for *HNF1B* with constellation of impaired glucose tolerance and kidney disease, or family history for these conditions. | Adult (n=18) and children (n=32) | MLPA deep sequencing of *HNF1B* exons validated by Sanger sequencing | 14/36 (38.9 %) | 36/50 (72%) |
| Rao et al., 2019 ^24^ | China | Prospective cohort  Multi-center | National Children Genetic Kidney Disease **Database** | Nephrologist  Molecular geneticist  Bioinformaticians  Clinical geneticist  Genetic counselors | Children with suspected genetic kidney disease | Children (n=1001) | Comprehensive testing using clinical WES with analysis of virtual panels | 421/1001 (42.1%) | NR |
| Shen et al., 2021 ^25^ | China | Retrospective cohort  Single center | **Multidisciplinary and sex-stratified approach for screening and management** of Children at risk of Fabry’s disease | Nephrologist, geneticist, genetic counsellor, cardiologist, gastroenterologist, neurologist, psychologist | Children at high risk of Fabry’s disease based on symptoms or positive family history | Children (n=35) | Sanger sequencing of the *GLA* gene | Dried blood spot triple-test screening of high-risk children: 5/35 (14.3%)  Universal newborn screening: 2/1420 (0.14%) | 35/35 (100%) |
| Miura et al., 2023 ^26^ | Japan | Retrospective cohort  Single center | **Genetic testing approach** for FSGS/SRNS | NA | Pediatric kidney transplant recipients whose kidney failure were due to FSGS/SRNS:  1. Familial/syndromic FSGS  2. Presumed primary FSGS  3. Undetermined | 23 families | Comprehensive testing using clinical WES with analysis of virtual panels | Familial/syndromic FSGS: 4/4 (100%)  Presumed primary FSGS: 0/8 (0%)  Undetermined FSGS: 10/11 (91%) | NR |
| Jung et al., 2023 ^27^ | Republic of Korea | Retrospective cohort  Single center | **Genetic testing** using index-only WES | Geneticist | Patients with suspected genetic kidney disease | Adult (n=23) and children (n=149) | Research WES confirmed by Sanger’s  CMA for CNV | 63/172 (36.6%) | 172/172 (100%) |
| Lim et al., 2024 ^28^ | Singapore | Prospective cohort  Multi-center | Pilot implementation of **nephrologist-led genetic service** for suspected monogenic glomerular disease and cost-effectiveness of genetic testing | Nephrologist  Clinical geneticist  Molecular geneticist  Genetic counselors | Patients with suspected genetic kidney disease including:  1. Glomerular disease  2. SRNS  3. CKD unexplained | NA | Clinically accredited targeted panel  Clinical WES | NA | NA |

*AUS = Australia; NZ = New Zealand; UK = United Kingdom; USA = United States of America; WES = Whole Exome Sequencing; WGS = Whole Genome Sequencing; CNV = Copy Number Variant; MLPA = Multiplex Ligation-dependent Probe Amplification; SRNS = Steroid-Resistant Nephrotic Syndrome; FSGS = Focal Segmental Glomerulosclerosis; ADPKD = Autosomal Dominant Polycystic Kidney Disease; CKD = Chronic Kidney Disease; NPH-RC = Nephronophthisis-related ciliopathies; aHUS = atypical Hemolytic Uremic Syndrome; CMA = Chromosomal Microarray Analysis; MDT = Multidisciplinary Team; HNF1B = Hepatocyte Nuclear Factor 1 Beta; GLA = Galactosidase Alpha; NA = Not Available; NR = Not Reported*

*Diagnostic yield (%) refers to the proportion of patients sequenced who received a genetic diagnosis. Test uptake rate (%) refers to the proportion of patients offered testing who underwent sequencing*

**Supplemental Table 7: Domain 4 (Stakeholder experience, n=4)**

| **Study (Author, Year)** | **Country** | **Study Design and Setting** | **Study Aims** | **Study Methodology** | **Stakeholder Groups** | **Stakeholder Reported Outcome Measures** |
| --- | --- | --- | --- | --- | --- | --- |
| Nevin et al., 2022 ^29^ | AUS | Retrospective mixed quantitative and qualitative  2 tertiary pediatric hospitals | To understand:  1. The experiences of families undergoing genetic testing;  2. The psychosocial impact of receiving a genetic test result; and  3. Parent information and support needs, regarding the delivery of genomic information by the nephrology team. | 1. Survey: Quality of Life Scale-Family Version (QoL-FV) tool  2. Semi-structured interview | 26 Parents of a child who had renal disease and had been referred for genetic testing and counselling | 1. Quality of life scale family version (QoL-FV) tool to assess the quality of life of caregivers and impact of parenting a child undergoing genetic testing.  7 items which includes:(all 7 items were ranked 1-10, 1=not at all to 10=extreme)  (a) Quality of life: mean 7.3  (b) Distress surrounding genetic testing: mean 2.5  (c) Distress undergoing genetic testing: mean 3.4  (d) Impact on personal relationships: mean 1.7  (e) Positive life changes: mean 3.5  (f) Uncertainty about the future: mean 7.2  (g) Distress at diagnosis and treatment: mean 8.6  2. Parents' experience in 4 domains:  (a) Psychosocial functioning at interview (QoL-FV)  (b) Experience of genetic testing process  (c) Psychosocial impact of a genetic test result  (d) The information needs and preferences of parents |
| Mallet et al., 2024 ^30^ | AUS | Editorial | To reflect on factors contributing to the early success of Australian Medicare Benefits Scheme genomic testings and the pathway ahead. | Australia national multi-center kidney genetics clinic network 10-year experience | Healthcare provider/researcher | Key barriers and facilitators of successful Australian Medicare Benefits Scheme genomic testings. |
| Kneifati-Hayek 2024 ^31^ | USA | Cross-sectional qualitative survey study  Multi-center | 1. To evaluate nephrologists’ knowledge, attitudes, and willingness to use genomic resources in clinical practice, and identify factors influencing their decision to order or refer patients for genetic testing.  2. To identify unmet needs among practicing US nephrologists to inform development of nephrology-tailored decision support tools. | Survey questions consisted of 6 sections:  1. Respondents' demographics and practice setting  2. Respondents' experiences with genomic resources  3. Attitudes toward using genomic resources in clinical care  4. Willingness to adopt new diagnostic tools  5. Barriers to using genomic resources  6. Knowledge and self-efficacy in using genomic resources | 319 United States based nephrologists who are board-certified and in active clinical practice. | 1. Demographics and practice setting characteristics  -74% had at least 5 years of attending-level experience in nephrology.  -Adult nephrologist (87%) vs pediatric nephrologist (13%)  75% spent at least 50% of their efforts in patient-facing care.  2. Experience using genomics  -Majority 76% had prior experience in ordering genetic testing.  -56% reported participating in returning genetic test results to patients.  -32% favored a clinical workflow in which nephrologists both ordered genetic testing for their patients and communicated the results.  3. Attitudes toward the utilization of genomic resources (5-point Likert scale)  (a) Clinical usefulness median score 4 (IQR 4-4))  (b) Training and preparedness median score 3 (IQR 3-3.5)  (c) Willingness to use new technologies median 20 (IQR 18-21) range 5-25  (d) Perceived self-efficacy median score 3(3-3.25)  (e) Perceived knowledge median score 3 (IQR 2.5-4)  (f) Objective knowledge median score 6 (IQR 5-6) |
| Fernandez et al., 2025 ^32^ | USA | Cross sectional qualitative online survey  Multi-center | To understand the views of paediatric nephrologists regarding genetic testing in clinical settings. | Online Survey about:  1. Understanding and experiences related to genetic testing in clinical settings, including the usefulness of genetic testing for clinical management, cascade testing, and procedures for returning results.  2. Barriers and possible facilitators to genetic testing. | 85 practicing paediatric nephrologists in USA | 1. Feasibility of use of genetic testing in daily clinical practice  -86% of nephrologists agreed that genetic testing fits into the process already in use for the care of kidney patients.  -56.5% agreed that they have enough time to facilitate the integration of genetic testing into clinical practice (56.5%)  - 68.2% can find/use reliable sources of information they need to apply genetics while caring for patients.  - 92.9% believe that genetic testing is relevant to their current clinical practice.  - 36.5% have a clearly designated person or teams lead the effort to incorporate genetic testing into clinical practice.  -58.8% agreed that a variety of strategies are being used to enable staff to use genetic testing.  2. Appropriateness, adoption, and acceptability of genetic testing  - Genetic testing was considered clinically important for disease diagnosis (92%), understanding (85%), prognosis (86%), treatment (84%), family counselling (88%), and kidney transplant planning (93%).  - The majority (70%) of participants would recommend genetic testing for family members, especially in the presence of a tailored lab report (91%). |

*AUS = Australia; USA = United States of America; QoL-FV = Quality of Life Scale – Family Version; IQR = Inter-quartile Range*

*The study by Nevin et al. also contributes to the domain of Ethical, Legal, and Social Implications (ELSI) in genomic testing, addressing psychosocial outcomes and parental information/support needs.*

**Supplemental Table 8: Domain 5 (Health economic and health system analysis, n=2)**

| **Study (Author, Year)** | **Country** | **Study Design and Setting** | **Study Aims** | **Study Methodology** | **Population** | **Outcomes** |
| --- | --- | --- | --- | --- | --- | --- |
| Meng et al., 2021 ^33^ | AUS | Prospective cohort  Multi-center | To elicit the willingness-to-pay (WTP) for genomic testing, using contingent valuation, among people with lived experience of genetic conditions in Australia | Parents of children with suspected genetic disorders completed a dynamic triple-bounded dichotomous choice (DC) contingent valuation survey. Adult patients or Parents of children with suspected genetic kidney disease or complex neurological and neurodegenerative conditions completed a payment card (PC) contingent valuation. | 360 parents of children with 6 categories suspected genetic disorders:  1. Mitochondrial disorders (N=24)  2. Developmental epileptic encephalopathy (N=44)  3. Leukodystrophy (N=13)  4. Malformations of cortical development (N=60)  5. Genetic kidney disease (N=159), and  6. Complex neurological and neurodegenerative conditions (N=60) | Cost benefit analysis:  1. Willingness-to-pay (WTP) for genomic testing  - mean WTP for genomic testing was estimated at AU$2830 (95% CI 2236-3424) for parents of children with mitochondrial disorders, epileptic encephalopathy, leukodystrophy, or malformations of cortical development genetic conditions based on the dichotomous choice data and AU$1914 (95% CI 1532-2296) for adults of parents of children with genetic kidney disease or complex neurological and neurodegenerative conditions based on the payment card data  - mean WTP across 6 cohorts ranged from lowest AU$1879 (genetic kidney disease) to highest AU$4554 (leukodystrophy) |
| Regnier et al., 2024 ^34^ | Multinational | Cross-sectional survey | To evaluate global access to diagnostic investigations (*CTNS* genetic testing, intra-leucocyte cystine) and cysteamine formulations in developing/transitioning economies (DEing/TrE) versus developed economies (DEed) and to document changes since 2011. | International cross-sectional survey of nephrology centres; 43-item Google-Forms questionnaire (demographics, genetics, biochemical monitoring, treatment availability & reimbursement, transition & multidisciplinary care) | Paediatric (94 %) and adult nephrology services in 109 centres across 49 countries caring for 741 patients with nephropathic cystinosis | 1. Access rates to genetics, intra-leucocyte cystine (IL-CL) testing, cysteamine (immediate & delayed release), eye-drops, dialysis, transplantation, multidisciplinary & transition programmes. Improvement since 2011 documented (e.g., genetics availability in DEing rose from 23 %→63 %; IL-CL 0 %→30 %).  2. Availability of *CTNS* gene sequencing (any method) captured: accessible in 63 % of DEing/TrE countries vs 100 % DEed;  3. Median reported cost USD 650 (DEing) vs 780 (DEed); reimbursement total in 37 % vs 79 % of countries, respectively. |

*AUS = Australia; WTP = Willingness-To-Pay; DEing/TrE = Developing or Transitioning Economies; DEed = Developed Economies; IL-CL = Intra-leucocyte Cystine Levels; CTNS = Cystinosin, Lysosomal Cystine Transporter (gene associated with nephropathic cystinosis)*

**Supplemental Table 9: Domain 6 (Kidney genomic implementation process evaluation, n=1)**

| **Study (Author, Year)** | **Country** | **Study Design and Setting** | **Study Aims** | **Study Methodology** | **Stakeholder Group** | **Outcomes** |
| --- | --- | --- | --- | --- | --- | --- |
| Best et al., 2023 ^35^ | AUS | Cross sectional  Multi-center | 1. To understand the process of carrying out the genomic intervention that are amenable for adaptation  2. To identify nongenetic physicians barriers to the implementation of genomic testing to inform future potential implementation strategy development. | Qualitative deductive semi-structured interviews using:  1. Process mapping  2. Theoretical Domains Framework (TDF) | 16 non-genetic clinicians were interviewed:  6 nephrologists, 4 neurologists, 1 cardiologist  11 in adult care and 5 in paediatric services. | 1. Process of genomic intervention that are amenable for adaptation  -identified 10 individual process map, 1 overview map, 16 common steps, and 9 steps amenable to adaptation.  - 6 were not explored at interview, and 1 was a patient decision-making step.  Phase 1: Ensuring appropriate patients receive genomic testing (TDF code: behavioural regulation)  Referral received and initial screening  Pre-clinic patient contacts to obtain family and pedigree history  Pre-clinic meeting to decide around eligibility, what test and how clinic will run  Patients seen in medical specialist genetic clinic  Patient consented.  Phase 2: Test ordering and interpreting variants  Appropriate information to labs  Clinical curation  Curated results to physician.  Phase 3: Providing results to patients (TDF code: Optimism)  Results to patients  2**.** Nongenetic physicians barriers to the implementation of genomic testing |

*AUS = Australia; TDF = Theoretical Domains Framework*

**Supplemental Table 10: List of global kidney genetics clinic leads collaborators (n=48)**

| **Full Name** | **Country** | **Affiliation** |
| --- | --- | --- |
| John Andrew Sayer | United Kingdom | Bioscience Institute, Newcastle University and The Newcastle upon Tyne Hospitals NHS Foundations Trust. |
| Richard Sandford | United Kingdom | University of Cambridge and Cambridge University Hospitals NHS Foundation Trust. |
| Jenny Patterson | United Kingdom | West of Scotland Regional Genomics Service. |
| Elizabeth Watson | United Kingdom | South West Genomics Laboratory Hub (NHS), Bristol. |
| Joanna Jarvis | United Kingdom | Department of Clinical Genetics, Birmingham Women’s and Children’s NHS Foundation Trust, Birmingham. |
| Helen M. Stuart | United Kingdom | Manchester Centre for Genomic Medicine, Manchester University NHS Foundation Trust, Manchester.  Division of Evolution, Infection and Genomics, School of Biological Sciences, Faculty of Biology, Medicine and Health, University of Manchester, Manchester. |
| Anjali Menon | United Kingdom | North Bristol NHS Foundation Trust. |
| Daniel P. Gale | United Kingdom | UCL Centre for Kidney and Bladder Health, University College London and National Registry of Rare Kidney Diseases, Bristol, UK. |
| Melanie MY Chan | United Kingdom | Medical Research Council Laboratory of Medical Sciences, Imperial College London.  Imperial College Healthcare NHS Trust, London. |
| Asheeta Gupta | United Kingdom | Department of Nephrology, Great Ormond Street Hospital, London.  Department of Translational Sciences, University of Bristol, Bristol. |
| Peter J Conlon | Ireland | Beaumont Hospital, Dublin 9 and Royal College of Surgeons in Ireland. |
| Aron Chakera | Australia | Renal Unit, Sir Charles Gairdner Hospital, Nedlands. |
| Kathy Nicholls | Australia | The Royal Melbourne Hospital and the University of Melbourne, Parkville. |
| Randall James Faull | Australia | Renal Unit, Royal Adelaide Hospital, University of Adelaide. |
| John Whitlam | Australia | Department of Nephrology, Austin Health, Heidelberg, Victoria.  Clinical Genetics Service, Austin Health, Heidelberg Victoria. |
| Michel Tchan | Australia | Dept of Genetic Medicine, Westmead Hospital, Sydney.  Sydney Medical School, Sydney University, Sydney. |
| Hugh J McCarthy | Australia | Child and Adolescent Health, Faculty of Medicine and Health, University of Sydney. |
| Parthasarathy Shanmugasundaram | Australia | Department of Renal Medicine, St George hospital, Kogarah, New South Wales. |
| Catherine Quinlan | Australia | Department of Nephrology, Royal Children's Hospital.  Kidney Regeneration, Murdoch Children’s Research Institute.  Department of Paediatric, University of Melbourne. |
| Zornitza Stark | Australia | Victorian Clinical Genetics Services, Murdoch Children's Research Institute, Flemington Road, Melbourne.  Australia and Department of Paediatrics, University of Melbourne, Melbourne. |
| Ella Wilkins | Australia | Victorian Clinical Genetics Services, Murdoch Children's Research Institute, Flemington Road, Melbourne.  Australia and Department of Paediatrics, University of Melbourne, Melbourne. |
| Andrew J. Mallett | Australia | Department of Renal Medicine, Townsville University Hospital, Townsville, Queensland. |
| Chirag Patel | Australia | Genetic Health Queensland, Royal Brisbane and Women’s Hospital, Herston, Queensland. |
| Jessica Ryan | Australia | Monash Health and Monash University. |
| Kushani Jayasinghe | Australia | Department of Nephrology, Monash Medical Centre, Melbourne, Victoria.  Monash University, Melbourne, Victoria.  Department of Clinical genetics, Melbourne Health. |
| Amali Mallawaarachchi | Australia | Department of Clinical Genetics, Royal Prince Alfred Hospital, Sydney.  Garvan Institute of Medical Research, Sydney. |
| Nicholas Cross | New Zealand | Department of Nephrology, Christchurch Hospital, Te Whatu Ora Waitaha Canterbury. |
| Ian Hayes | New Zealand | Genetic Health Service New Zealand-Northern Hub, Auckland. |
| Whitney Besse | United States | Department of Internal Medicine, Section of Nephrology, Yale School of Medicine, New Haven, Connecticut. |
| Xiangling Wang | United States | Departments of Kidney Medicine, and Medical Genetics and Genomics, Cleveland Clinic, Cleveland, Ohio. |
| Jordan G. Nestor | United States | Department of Medicine, Division of Nephrology, Columbia University, New York. |
| Amar J. Majmundar | United States | Department of Pediatrics, Boston Children’s Hospital, Boston, Massachusetts. |
| Marie C Hogan | United States | Division of Nephrology and Hypertension, Mayo Clinic, Rochester, Minnesota. |
| Christie P Thomas | United States | Department of Internal Medicine, University of Iowa, Iowa City, Iowa. |
| Michael Brendan Shannon | United States | University of Washington Harborview Medical Center, Seattle, Washington. |
| Ana C. Onuchic-Whitford | United States | Brigham and Women's Hospital, Renal Division, Boston, Massachusetts. |
| Andrew L. Lundquist | United States | Massachusetts General Hospital, Division of Nephrology, Boston, Massachusetts. |
| Dervla M Connaughton | Canada | Department of Biochemistry, Schulich School of Medicine & Dentistry, University of Western Ontario, London, Ontario.  Division of Nephrology, Department of Medicine, London Health Sciences Centre, London, Ontario. |
| Moumita Barua | Canada | Division of Nephrology, University Health Network, Toronto, Ontario, Canada.  Department of Medicine, University of Toronto, Toronto, Ontario, Canada. |
| Roser Torra | Spain | Fundacio Puigvert, IR Sant Pau, RICORS2040 renal, Barcelona, Spain. |
| Isabel Galán Carrillo | Spain | Reina Sofía Hospital (Murcia)  University Complutense Madrid  Murcia University |
| Sandrine Lemoine | France | Hospices Civils de Lyon, Nephrology, Dialysis and Renal Physiology Department, Centre De Référence Maladies Rénales Rares MAREGE. |
| Laurent Mesnard | France | Soins Intensifs Néphrologiques et Rein Aigu (SINRA), Nephrology Department, Tenon Hospital, Assistance Publique – Hôpitaux de Paris, Paris, France.  European Rare Kidney Disease Reference Network (ERKNet).  Inserm UMR_S1155, Paris. |
| Albertien M. van Eerde | The Netherlands | Department of Genetics, University Medical Center Utrecht, Utrecht. |
| Jan Halbritter | Germany | Department of Nephrology and Medical Intensive Care, Charité Universitätsmedizin Berlin, Berlin. |
| Asaf Vivante | Israel | Sheba Medical Center, Nephrogenetic Clinic, Tel Aviv University. |
| Janewit Wongboonsin | Thailand | Renal Division, Department of Internal Medicine, Faculty of Medicine Siriraj Hospital, Mahidol University, Bangkok, Thailand.  Bumrungrad Genomic Medicine Institute and Department of Medicine, Bumrungrad International Hospital, Bangkok, Thailand.  Division of Renal Medicine, Brigham and Women's Hospital, Boston, Massachusetts, United States.  Department of Medicine, Harvard Medical School, Boston, MA, United States. |
| Becky Mingyao Ma | Hong Kong SAR, China | Division of Nephrology, Department of Medicine, Queen Mary Hospital, The University of Hong Kong, Hong Kong, P.R. China. |

*A total of 48 survey responses were received, representing 48 unique respondents. One respondent completed two separate surveys corresponding to two distinct centres where she is concurrently employed. In addition, two respondents jointly completed a single survey on behalf of one conjoint clinic that they co-run.*

**Supplemental Figure Legends**

**Supplemental Figure 1. PRISMA flow diagram of study selection process**
A total of 4,375 records were identified through database searches: MEDLINE (n = 1,979), Scopus (n = 1,638), Embase (n = 763), and Google Scholar (n = 4). After removal of 746 duplicates, 3,629 records were screened by title and abstract, resulting in 148 full-text articles assessed for eligibility. Of these, 88 studies were excluded for reasons including wrong setting (n = 24), editorial/review articles (n = 10), wrong indication (n = 1), wrong intervention (n = 2), wrong study design (n = 3), no clinic model reported (n = 40), or abstract only with no full-text publication (n = 8). Sixty studies were included in the final scoping review.

**Supplemental Figure 2.** **Annual and regional distribution of included publications (n = 60)**
Stacked bar chart showing the number of publications from 2013 to 2025, stratified by region and color coded by geographic location: Asia (yellow), Australia/New Zealand (blue), Europe (green), North America (red), and UK/Ireland (purple). Publication output increased over time, with peaks observed in 2021 and 2024 (12 publications each).

**Supplemental Figure 3. Distribution of publications by key thematic domains (n = 60)**
Publications were coded to one or more domains based on study focus. The most frequently represented domains were: genomic utility in kidney disease outside of clinic settings (navy, n = 26), clinic model description and evaluation (green, n = 25), and methodology or infrastructure development for genomic implementation (yellow, n = 33). Fewer studies addressed stakeholder experience (orange, n = 7), implementation process evaluation (light purple, n = 2), health system or economic analysis (dark purple, n = 2), and ethical, legal, or social implications (red, n = 1). Total counts exceed 60 due to overlapping classification.

**Supplemental Figure 4.** **Domain classification of included studies by first author and region (n = 60)**
Each bar represents one publication, grouped by region and labeled by first author. Studies were coded to one or more thematic domains, with color coding consistent with Figure 4: green (clinic model description and evaluation), navy (genomic utility in kidney disease), yellow (methodology or infrastructure development), orange (stakeholder experience), light purple (implementation process evaluation), dark purple (health system or economic analysis), and red (ethical, legal, or social implications). Bar height reflects the number of domains assigned per study (range: 1–3), illustrating the multidimensional focus of some publications.

**Supplemental Figure 5a.** **Professional background of survey respondents by region**
Bar chart showing the distribution of professional roles among survey respondents (n = 48), stratified by region. Roles are color coded as follows: adult nephrologist (dark blue), pediatric nephrologist (orange), clinical geneticist (dark green), genetic counselor (purple), nephrologist-geneticist (light blue), and laboratory geneticist (light green). Adult nephrologists were the most common respondents across all regions.

**Supplemental Figure 5b.** **Years of professional experience among survey respondents (n = 48)**
Pie chart illustrating respondents’ self-reported years of experience working in kidney genetics or related fields. Most had 10 years or more (green, 44%) or 7–9 years (pink, 35%) of experience, followed by 4–6 years (blue, 15%) and 1–3 years (yellow, 6%).

**Supplemental Figure 6.** **Types of genetic testing laboratories used in kidney genetics clinics, stratified by region (survey respondents, n = 48)**
Sunburst chart showing the distribution of laboratory types used for genetic testing across five regions. The inner ring represents region: Australia/New Zealand (AUS/NZ, blue), Asia (orange), Europe (green), North America (red), and UK/Ireland (purple). The outer ring displays the types of laboratories utilized, including in-house (clinically accredited lab within the same institution), commercial (external accredited labs), national laboratories (regionally funded accredited services), research (non-accredited labs requiring reflex confirmation), and hybrid models. Hybrid refers to combinations involving in-house, commercial, and/or research laboratories.

UK/Ireland and Europe primarily used in-house and national labs; AUS/NZ used a mix of in-house, commercial, and hybrid approaches. North America predominantly used commercial and hybrid labs, while Asia reported smaller but varied combinations of in-house, commercial, and research testing models.

**Supplemental Figure 7.** **Sources of funding for genetic testing in kidney genetics clinics, stratified by region (survey respondents, n = 48)**
Sunburst chart showing the reported funding mechanisms specifically for genetic testing across five regions. The inner ring represents region: Australia/New Zealand (AUS/NZ, blue), Asia (orange), Europe (green), North America (red), and UK/Ireland (purple). The outer ring categorizes funding sources as: public (government or health system–funded), insurance/out-of-pocket, public/insurance, mixed (combination of public, private, research, or philanthropic sources), and research (non-clinically funded academic or grant-based testing). Public funding for genetic testing was the predominant model in UK/Ireland (92%), Australia/New Zealand (81%), and Europe (57%). In contrast, North America reported a more fragmented landscape, with reliance on insurance/out-of-pocket (36%) and mixed funding models (27%). Asia showed limited but mixed sources of funding.
